# Provides Apparent Predictive Biomarker in a Phase I/II Study of Pembrolizumab With Gemcitabine in Patients with Previously-Treated Advanced Non-Small Cell Lung Cancer (NSCLC)

**DOI:** 10.1101/2024.12.20.24317919

**Authors:** Rachel E. Sanborn, Shawn M. Jensen, Christopher Paustian, Shu-Ching Chang, Eloy Vetto, Quyen Vu, Venkatesh Rajamanickam, Brady Bernard, Yoshinobu Koguchi, William Redmond, Bernard A. Fox

## Abstract

Retrospective characterization of cell-cell relationships in the tumor microenvironment provides significantly better predictive power than PD-L1 expression, tumor mutational burden (TMB), or gene expression profiles. In this small study assessing the safety and possible efficacy of gemcitabine and pembrolizumab in immunotherapy-naïve patients with NSCLC who have received prior treatment, we investigated both standard and novel immune parameters on 16 enrolled patients. The combination of gemcitabine and pembrolizumab could be administered safely but did not demonstrate synergism compared with historical controls. Novel findings of this study are that elevated frequencies of Tregulatory cells near CD3 T cells at baseline was associated with improved outcome to treatment (p<0.05). Integrating this Treg / T cell relationship metric together with overall T cell density yielded a tumor inflammation score which correlated (p<0.002) with disease response. We postulate this is indicative of ongoing anti-cancer immune response. Additionally, while prior studies documented that IgG Ab responses to TAA can identify targets of a coordinated T and B cell response and evidence of immune surveillance, this study found that high autoantibody responses, while not statistically significant, trended towards a worse outcome (p=0.06). This suggests to us that tumors may have developed mechanisms to escape the immune response to these TAAs.

## Introduction

Non-small cell lung cancer (NSCLC) is the most common cancer killer ^1,2^. Checkpoint inhibitors have significantly improved survival for patients with advanced NSCLC ^3–6^, although the benefit is limited for the majority of patients ^5,7–10^, and improved therapeutic approaches are needed.

Gemcitabine is an active agent in NSCLC. Single agent gemcitabine has progression-free survival (PFS) ranging between 2.5-4.25 months in advanced NSCLC, and median survival of 4.4-7 months in the first-line setting for patients over the age of 70 ^11–13^. Phase III trials of gemcitabine in the second-line setting for advanced NSCLC have not been performed, but in phase II studies the median time to progression is 2.5 months, with median overall survival of 9.5 months, and overall response rate is 18.5% ^14^. Although gemcitabine historically has been considered to be purely immunosuppressive, preclinical studies have indicated that gemcitabine increases antigen cross-presentation, as well as increases the percentage of intratumoral primed tumor-specific CD8 T cells, without increasing functional tolerance ^15^. Gemcitabine has also been reported to preferentially reduce regulatory T cells (Tregs), providing additional rationale for combining with checkpoint inhibition^16^.

Despite the evaluation of multiple chemotherapy agents with checkpoint inhibitors, pembrolizumab and gemcitabine had not previously been studied in combination. This study was proposed as a feasibility trial evaluating the combination of pembrolizumab and gemcitabine for immunotherapy-naive patients with previously-treated advanced NSCLC. To inform about the immune status of patients prior to and during treatment, flow cytometric analyses of peripheral blood cell populations, and phage display immunoprecipitation (PhIP) of sera were performed. Additionally, multiplex immunoflouresence (mIF) analyses of the tumor were performed on pre-treatment biopsies. Based on the work of Lu et al., we were particularly interested in evaluating the potential predictive power that characterization of cell-cell relationships in the pre-treatment biopsy might provide ^17^.

## Materials and Methods

### Patients

Key eligibility criteria allowed for patients with advanced NSCLC treated with one to three prior lines of systemic therapy. Patients were required to have good performance status (Eastern Cooperative Oncology Group status 0 or 1), and adequate organ function as outlined in the clinical protocol (Supplementary Figure 1). Measurable disease by Response Evaluation Criteria in Solid Tumors (RECIST) version 1.1 was required^18^. Treated stable brain metastases were allowed.

Key exclusion criteria included prior exposure to gemcitabine or checkpoint inhibitors, any elements of small cell carcinoma, history of pneumonitis requiring steroid intervention or interstitial lung disease, and otherwise as listed in the protocol. Trial design, assessments and toxicities are included in supplemental data.

### Correlative studies

#### PD-L1 testing

PD-L1 expression was evaluated utilizing the 22C3 assay.

#### Flow cytometry

Whole blood immune profiling assays were conducted as previously described ^19,20^. Briefly, heparinized whole blood was stained with a cocktail of antibodies to identify Tregs as well as monocytic myeloid-derived suppressor cells (Mo-MDSCs). Samples were acquired with a BD LSRFortessa (BD Biosciences). Data were analyzed with FlowJo version 10 (BD Biosciences). Tregs were defined as CD3^+^CD4^+^CCR4^hi^CD127^low^CD25^hi^ cells (Fig. S2). Monocytic MDSCs were defined as lineage (CD3, CD7, CD19, and CD20) negative, CD15^−^CD11b^+^CD33^+^CD14^+^HLA-DR^low^ cells (Fig. S3).

#### Phage Display Immunoprecipitation Sequencing (PhIP-Seq) and Data Analysis

PhIP-Seq was performed by CDI Labs as previously described on sera from patients at times specified ^21^.

#### mIF Analysis of Tumor Biopsies

A five-plex mIF panel (PD-L1, Cell Signaling Technologies E1L3N; PD-1, Abcam EPR4877(2); CD3, Abcam SP7; FOXP3, Abcam 236A/E7; and CK, Novous Biologicals AE1/AE3) developed and optimized, and shown to have quantitative equivalence to immunohistochemistry (IHC) chromogenic assays was used to assess FFPE sections where available^22^. Reagents described previously were used to stain samples on a Leica Bond Rx autostainer. 20X Digital images were captured with the Akoya Vectra-Polaris multispectral imaging platform and analyzed using InForm Software (Akoya Biosciences). The total number of cells per mm^2^ was enumerated for all the examined cell phenotypes and statistical analysis between groups was established using Graphpad Prism. Spatial relationship analysis between cell phenotypes was performed using phenoptr Reports (Akoya Biosciences). Tumor Inflammation Score was determined by multiplying (the average number of T_reg_ cells within 15 mm of a CD3^+^ T cell) X (the CD3^+^ T cell density for that tissue section).

#### Statistical analysis

The primary outcome for analysis was frequency of toxicities with the combination of gemcitabine and pembrolizumab. Secondary outcomes included PFS, OS, and ORR. Exploratory analyses included association between immune response with serial protein array measures from peripheral blood samples and ORR, PFS, and OS; association between immune response times of interest and PFS and OS; association between baseline PD-L1 expression in tumor tissue with PFS, OS, and best overall response; and association between baseline tumor tissue inflammation score and PFS, OS, and best overall response. All exploratory analyses were assessed as retrospective analysis.

Cox proportional hazards regression of PFS and OS were used to assess univariate effects of baseline PD-L1 expression and tumor inflammation score measures. Multivariate survival Cox proportional hazards models were used to test association of PD-L1 and tumor inflammation score measures while controlling for each other with PFS and OS. T-tests, or Wilcoxian rank-sum tests if nonparametrics were needed, were used to test the effects of the baseline PD-L1 expression, tumor inflammation score, and humoral immunity to possible TAA as measures on best overall response. Log transformations were used as needed to meet model assumptions. (Consort diagram, Figure S4)

## Results

### Patients

Sixteen patients were enrolled between July 2015, and August, 2018. Baseline patient characteristics are summarized in Table S1. The median age was 64 years (range, 53 to 75). The majority of patients were Caucasian/Non-Hispanic (14), had adenocarcinoma as primary histology (12), and ECOG status 1 (14 patients). The majority of patients had received one line of prior therapy (12 patients) primarily for treatment in the metastatic setting, although three patients were enrolled after progression after prior definitive concurrent chemoradiation for stage III disease.

PD-L1 status was available for 14 patients (two patients had insufficient tissue for results). PD-L1 status was high (≥50%) for three patients, four had PD-L1 status 1-49%, and seven had no PD-L1 expression detected.

### Efficacy

The median PFS (Figure 1A) and OS (Figure 1D) for the 16 patients was 3.10 (95% CI 2.60-5.68) and 8.20 (95% CI 5.03-28.29) months, respectively. The median duration of therapy was 2.2 months (range, 1.6-4.4 months).

**Figure 1:**
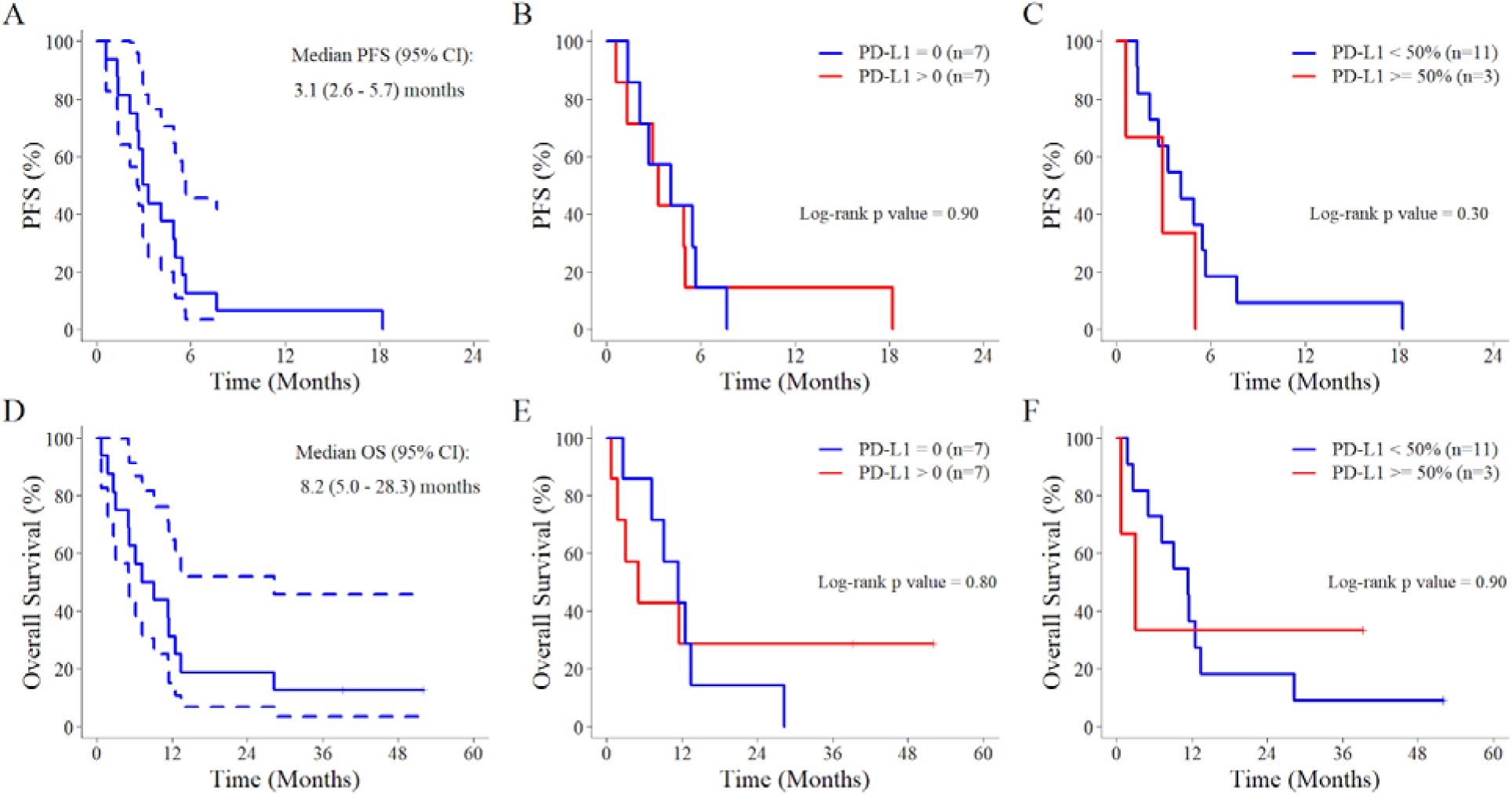
The median PFS for the 16 patients (panel A), and then separated by expression of PD-L1 (panels B and C). The median OS for the 16 patients (panel D), and then separated by expression of PD-L1 (panels E and F).

Two patients were still alive at the time of manuscript writing. One of these patients had discontinued therapy after developing grade 3 pneumonitis after cycle 3. This patient has been monitored expectantly with active surveillance since, requiring a single course of radiation to a solitary growing lung nodule, and has not yet required further systemic therapy (>3 years later). The other patient who remains alive experienced confirmed disease progression after cycle 24 pembrolizumab (best response was stable disease) and transitioned to subsequent systemic therapy. Of note, this patient was identified as having a BRAF V600E mutation, and subsequently was treated with combination tyrosine kinase inhibitors, as well as further systemic therapies.

Sixteen patients were evaluable for response. Partial response was identified as the best overall response in only two patients (ORR 12.5%), with no complete responses. Nine patients experienced stable disease (disease control rate, 68.8%). Figure S5 illustrates a spider plot of response.

### Correlative outcomes

#### Outcomes vs PD-L1 expression

No differences were seen in PFS or OS in patients with PD-L1 positive expression compared with PD-L1 negative tumors (Figure 1B and 1E, respectively). Additionally, no differences in PFS or OS were identified with using a cutpoint of 50% for PD-L1 expression levels (Figure 1C and 1F, respectively). Figure S6 delineates survival based upon different levels of PD-L1 expression. The patient with PD-L1 expression level of 100% had experienced Grade 3 pneumonitis as the reason for study discontinuation, and remains under active surveillance without further systemic therapy greater than three years later. The other patient who remains alive (>4 years later) had PD-L1 expression level of 40%.

Based upon the limited sample numbers, the above findings are hypothesis-generating only, and not statistically significant.

#### Peripheral blood immune profiling

It was reported that NSCLC patients with extrathoracic metastasis have elevated Mo-MDSCs in peripheral blood, which correlated with poor response to chemotherapy^23^. Therefore, we examined whether patients who did not respond to therapy exhibited higher frequency of Mo-MDSCs in the peripheral blood at baseline than those who responded to treatment. We also looked at Tregs as another suppressive immune cell population. Although we did not see any difference in peripheral blood Tregs (Fig. 2A), we observed a higher frequency of Mo-MDSCs in the peripheral blood at baseline in patients who did not respond to therapy (progressive disease or stable disease followed by progression) than those who did respond (partial response or sustained stable disease) (Fig. 2B). Representative flow plots from patients with progressive disease (Fig. 2C) or partial response (Fig. 2D) are shown. We also observed that peripheral blood Mo-MDSCs frequency was negatively associated with PFS (Fig. 2E).

**Figure 2.**
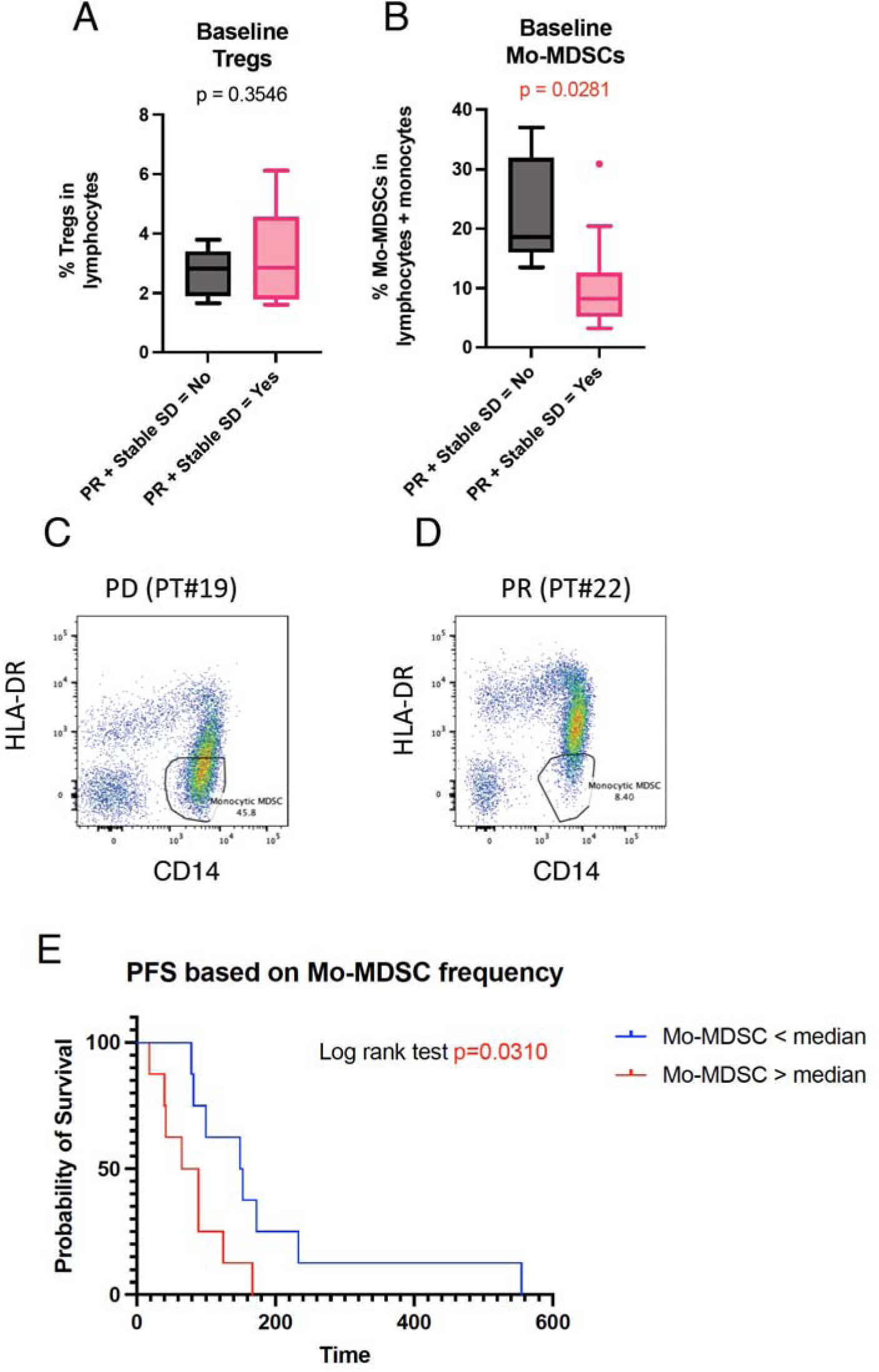
A higher frequency of Mo-MDSC at baseline correlated with poorer clinical outcomes. (A, B) Whole blood flow cytometric analysis for frequency of Tregs (A: CD45+CD3+CD4+CCR4+CD127lowCD25hi) or Mo-MDSCs (B: CD45+Lineage-CD14+CD33+CD11b+HLA-DRlow) in peripheral blood. Patients with (n=10) or without (n=6) progression were compared. Unpaired t test with Welch’s correction was used for statistical evaluation. (C, D) Representative flow cytometric data for Mo-MDSCs from a patient with PD (C) or PR (D). FACS plots from CD45+Lineage-CD33+CD11b+ between patients with a higher (Red: Mo-MDSC > 11.76%) and lower (Blue: Mo-MDSC < 11.76%) frequency of Mo-MDSC in peripheral blood at baseline. Log rank test was used for statistical evaluation.

#### Assessment of pre-existing and on treatment antibody responses

Tripathi and colleagues reported that patients with newly diagnosed NSCLC had antibody responses to self antigens, TAA, that correlated with cytotoxic T cell responses to epitopes of the same antigen presented on HLA-matched NSCLC cell lines ^24^. Based on these observations we evaluated whether the existence or development of new antibody responses correlated with response to treatment. All 16 patients provided baseline sera for PhIP-Seq interrogation. These sera reacted to as few as 6 to as many as 310 autoantigens, with the mean and median at 109 and 60 respectively. As seen in Figure 3A, no significant correlation between baseline PhIP-Seq hits and best response was evident. For any given hit the reactivity was assessed through a three-part data analysis pipeline to ensure robust qualification of hit calls.

**Figure 3.**
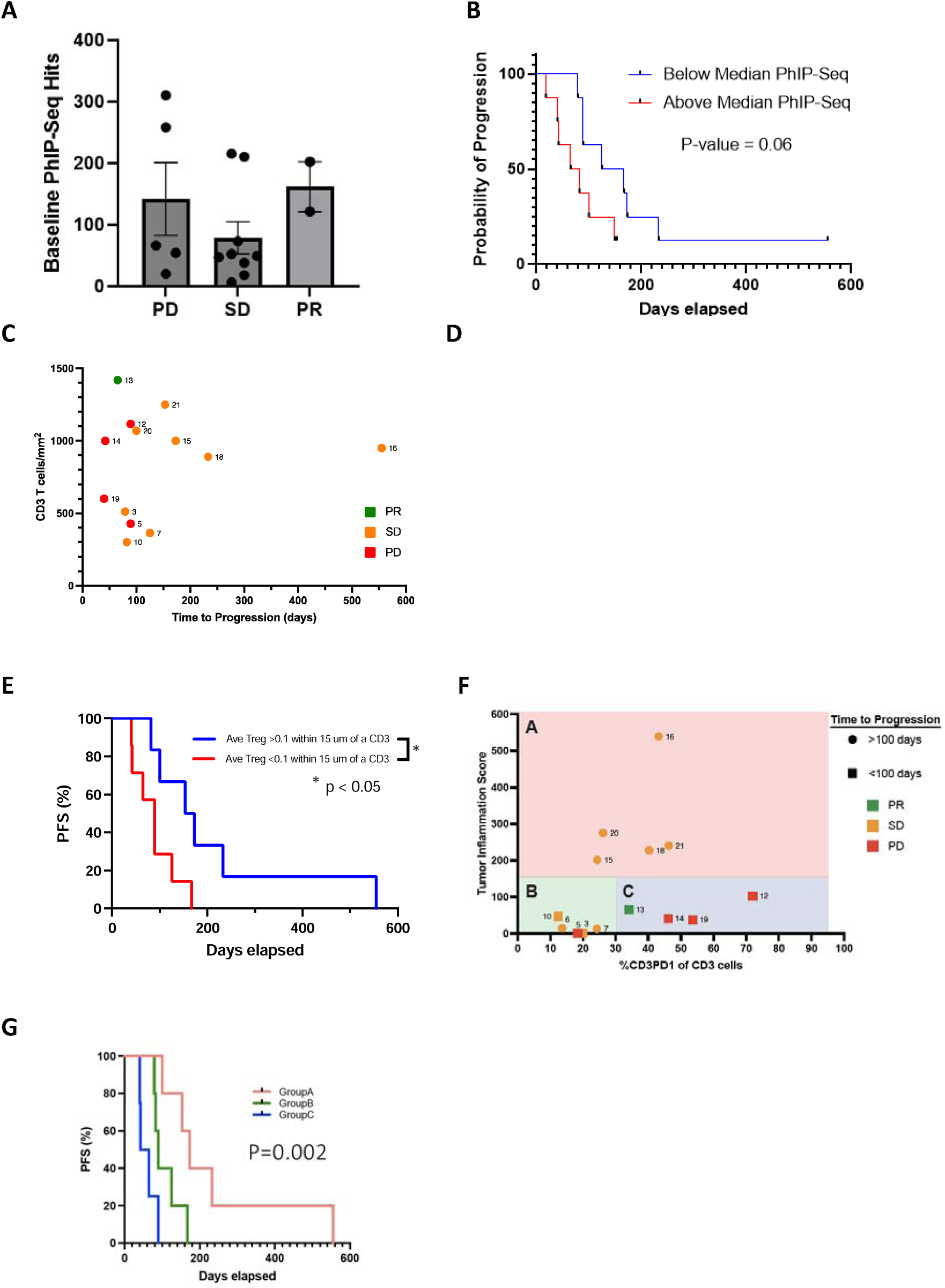
Immune correlates and outcomes. A) Number of hits to individual self-antigens shown (Filled circles). PD = Progressive Disease, SD = Stable Disease, PR = Partial Response. Histogram and error bars represent mean and SEM respectively. B) The probability of disease progression for patients with greater than (Red) or less than (Blue) the median number of PhIP-Seq hits at baseline. C) The CD3 T cell density (cells/mm_2_) plotted for the time to progression for each patient. D) The average number of regulatory T cells (T_reg_, CD3+FoxP3+) within 15um of a CD3 cell plotted for the time to progression for each patient (¡ = >100 days;” “ < 100 days). E) Patient time to progression curves plotted for groups with greater than 0.1 T_reg_ cells on average within 15um of a CD3 cell (Blue) versus less than 0.1 regulatory T cells on average within 15um of a CD3 cell (Red). F) Tumor Inflammation Score (Average number of T_reg_ cells within 15um of CD3 cells X CD3 T cell density) versus % of CD3 cells expressing PD-1 (¡= >100 days progression-free;” = < 100 days progression-free). G) Patient time to progression curves plotted for 3 groups segregated by Tumor Inflammation Score by %CD3PD1+ (shown in Figure 3F).

The median number of hits at baseline was 60. Using that as a cut-off, Kaplan-Meier curves comparing survival or disease progression were generated. Patients making greater than median auto-antibody responses had shorter time to disease progression and survival, however, these trends were not statistically significant (Figure 3B). Six patients were able to donate samples at the C6D1 timepoint, where reactivity to 12 to 587 hits were recorded. Responses to antigens that were not targeted at baseline are shown in Figure S7A. Reactivities ranged from as few as 4 new responses to as many as 412 new responses to 405 different self-antigens for the patient identified as “PD1G20”. This indicates that this patient generated autoantibodies against 405 proteins, with several responses against multiple epitopes within a protein. For a more quantitative assessment of hits, Z-scores were calculated comparing how many SD from the beads-only control the sample was. A handful of responses stood out as more than 50 SD from the control (Figure S7B).

#### Evaluation of tumor immune infiltrates and development of tumor inflammation score

To explore whether the immune contexture present in the tumor prior to treatment correlated with response, we undertook mIF analysis of biopsies for 14 patients where tissue was available. The frequency of CD3^+^ T cells in the tumor and associated stroma were enumerated and included an evaluation of FoxP3 expression and whether T cells were PD-1^+^ or PD-1^-^. A summary of findings are presented in Figure S8. Evaluation of T cell subsets and PFS and OS showed trends, particularly for patients with higher percentages of PD-1^+^ CD3 T cells tending to fare worse, but none of these evaluations reached statistical significance (Figure 3C & Figure S9). We next examined the relationship of T_reg_ cells to CD3^+^ T cells, by evaluating the number of T_reg_ cells within 15 or 30 µm of a CD3^+^ T cells. The average number of T_reg_ cells within 15 mm of a CD3 cell versus time to progression is shown in Figure 3D, and an association between higher numbers of T_reg_ cells around CD3^+^ T cells in the pretreatment biopsy and increased survival was identified (Figure 3E). Next, we posited that this increased frequency of T_reg_ cells within close proximity to CD3^+^ T cells could be a marker of an ongoing in situ anti-cancer immune response requiring Treg cells to control that response. So we combined that measurement with the overall CD3^+^ T cell density to obtain a “Tumor Inflammation Score” (TIS). Plotting the TIS against the percentage of CD3^+^ cells that were PD-1^+^separated the patients into three groups (Figure 3F). Patients with a high TIS were found in group A (Figure 3F, Panel A), patients with a low TIS and with less than 30% PD-1^+^ CD3^+^ T cells were in group B (Figure 3F, panel B), and patients with a low TIS and a high percentage of PD-1^+^ CD3^+^ T cells were in group C (Figure 3F, panel C). Group B contained patients with mixed outcomes (4 SD and 1 PD) and group C had 3 patients with progressive disease and 1 patient with a partial response. Pairwise comparisons using Log-rank test showed that there was significant difference in PFS (Figure 3G) between Groups A and C (p=0.008), but marginally significant between Groups A and B (p=0.05) and Groups B and C (p=0.068), based on Bonferroni-Holm method of adjustment. While the numbers in this study are small, plotting progression-free survival for the patients stratified into these three groups identified a significant difference (p=0.002, Figure 3G).

## Discussion

In this study, the combination of gemcitabine and pembrolizumab was feasible, without unanticipated significant toxicity. No synergistic signal was seen, however, with median PFS of 3.10 months; not different than the protocol-specified historical control of single-agent gemcitabine^11–13^. Median OS was 8.2 months; not improved compared with the previously published historical control of median OS 9.5 months with gemcitabine alone ^11–13^. The ORR of 12.5% was disappointing, given historical controls of ORR 18.5% with gemcitabine alone ^14^.

The PROLUNG study was a randomized phase II trial comparing pembrolizumab plus docetaxel to docetaxel in patients with previously treated advanced NSCLC^25^. This study evaluated a similar patient population, and included patients with tumors that had negative PD-L1 expression.

Pembrolizumab with docetaxel demonstrated improvement in ORR compared with docetaxel (42.5% vs 15.8%). Improvement in PFS was also seen with the combination (9.5 months vs 3.9 months). The PROLUNG trial results imply a potential synergism of pembrolizumab with docetaxel in previously-treated NSCLC, which was not demonstrated with gemcitabine and pembrolizumab in our trial.

Since our study, PD-1 inhibitors have been evaluated in combination with gemcitabine and platinum chemotherapy in patients with previously-untreated advanced squamous cell lung cancer, confirming feasibility, although there has not been separate discussion of relative synergistic activity ^6,26,27^. The POSEIDON trial, a randomized phase III study of durvalumab, tremilimumab, and platinum-doublet chemotherapy in the first-line setting for advanced NSCLC, indicated in an exploratory analysis concern for possible diminished benefit with a gemcitabine-based doublet compared with other platinum doublets, with the acknowledgement of the paucity of published data specifically pertaining to gemcitabine/immunotherapy combinations in NSCLC^28^.

Our results did not demonstrate a signal of differential outcomes for patients with high (≥50%) compared with lower (<50%) PD-L1 expression when treated with gemcitabine and pembrolizumab. Conclusions about implications of the combination effect, however, cannot be made due to the extremely small sample size.

IgG antibody responses to a specific protein are well established indicators of a CD4 T cell response to an epitope of the same antigen, as Ig class switching of B cells is generally attributed to contact-dependent and independent interactions with CD4 T cells ^29^. Correlative B cell and T cell responses to an epitope of the same antigen are not limited to CD4 T cells. Tripathi and colleagues reported that sera from patients with newly diagnosed NSCLC contained IgG against a large number of self proteins, for which T cell epitopes of those non-mutated proteins were presented by HLA of NSCLC cell lines^24^. They identified that patients with IgG responses to specific proteins also had cytotoxic CD8 T cell responses against these non-mutated TAA that were found to be shared by NSCLC, and could lyse HLA-matched NSCLC cell lines that naturally presented the TAA peptide. These findings provide evidence of immune surveillance against NSCLC in patients that are newly diagnosed and yet to have started treatment. Employing a preclinical model, our group demonstrated that vaccination with a complex cancer vaccine containing more than 50 self-proteins and 50 potential neoantigens generated a coordinated IgG and CD8 T cell response against an epitope contained in a 15 amino acid sequence of the specific antigen^30^. In an unbiased screen, some CD8 T cell responses identified by an IgG response were stronger to the native wild type epitope, while others were stronger to the neoantigen^30^.

Based on the above noted studies, we hypothesized that the induction of an antibody response to self-proteins, that are potential TAAs, could be an immune correlate to the generation of an anti-tumor T-cell response. By interrogating patient sera with peptides stemming from 24,239 individual open reading frames (ORFs) expressed on the surface of T7 Bacteriophage, we took a near exhaustive look for these hypothesized antibody responses among canonical proteins. One caveat of these experiments is that the 90mer peptides presented on the bacteriophage are limited in their ability to attain their natural tertiary conformation. Likewise, quaternary structures were not available for testing. Another difference between our work and that of Tripathi et al., was that their report was characterizing antibody responses of newly diagnosed patients with NSCLC, not patients following 1 or more lines of treatment. Given the small size of this study, we could not correlate a therapeutic benefit with the induction of an antibody response to self-proteins. We did see a correlation between a higher number of autoantibodies and poor survival and disease progression. While these associations were not statistically significant, they may suggest that patients had made a broad immune response (both B cell and T cell immune responses) to numerous antigens. We hypothesize this may have led to induction of immune escape mechanisms, including loss of HLA or exhaustion of tumor-reactive T cells that were primed to available cancer antigens. This is seen as consistent with patients who progressed more rapidly having a higher number of antibody responses, potentially suggesting that they had already utilized the spectrum of antigens that their T cells could recognize and those T cells were terminally exhausted, or their tumors had downregulated expression of those antigens. In the case where a patient’s tumor-reactive T cells are exhausted, the identification of noncanonical cancer antigens (that because of their short-lived nature are not cross-presented and require priming) could provide a wave of new T cells that in the context of combination immunotherapy may recover therapeutic immunity ^31,32^. This is a strategy our group is already exploring in a different combination immunotherapy trial.

Our flow-cytometry data confirmed the findings of an earlier study: metastatic NSCLC patients respond poorly to immunotherapy when they have a higher frequency of Mo-MDSCs in the periphery^23^. Our cohort showed no evidence that this form of immune suppression was overcome by immunotherapy with anti-PD-1. Importantly, peripheral enumeration of Treg cells did not correlate with outcomes, a finding that contrasts with our interrogation of the TME.

Evaluation of the TME using mIF identified a correlation between the presence of FoxP3^+^ T regulatory cells in proximity to CD3^+^ T cells and progression-free survival. We speculate that the presence of FoxP3^+^ T cells indicates a mechanism to control an ongoing anti-cancer T cell response. Combining this metric with T-cell infiltration into the tumor environment (T-cell density) allowed us to define a Tumor Inflammation Score (TIS). We further hypothesize that the administration of gemcitabine, an agent reported to eliminate FoxP3^+^ T_reg_ cells, in this trial could reduce and/or eliminate the regulatory component leading to an increased anti-cancer activity of the resident CD3^+^ T cells. Such activation of the endogenous immune response could lead to expression of PD-1, cytokine release in the TME, and upregulation of PD-L1 expression. Using the Tumor Inflammation Score and the percentage of CD3 T cells expressing PD-1, we identified a potential novel biomarker panel that, based on pre-treatment

biopsies, could be used to sort patients into 3 compartments. These compartments provide possible insights that may be used to personalize therapy in the future. Further, these findings underscore the importance of obtaining biopsies of patients enrolled onto clinical trials as we seek to personalize therapies with the goal of improving response rates.

## Declarations

The protocol was reviewed and approved by the Institutional Review Board at Providence Portland Medical Center, and was conducted in accordance with the Declaration of Helsinki. All patients provided written informed consent.

## Availability of data and material

Data are available on reasonable request.

## Competing interests

R. E. Sanborn reports: Grant funding for investigator-sponsored trial from AstraZeneca, Merck;

Consulting fees: GlaxoSmithKline, AstraZeneca, Janssen Oncology, Macrogenics, Daiichi Sankyo, Sanofi,

BeiGene, Gilead, Regeneron, Targeted Oncology, G1 Therapeutics, GE HealthCare, Amgen, Lilly

Oncology; Honoraria for educational presentations, manuscripts: EMD Serono, Illumina, GameOn!,

OncLive, Binay Foundation, APP Oncology, Masters in Thoracic Oncology

S. M. Jensen reports grants from Bristol Myers Squibb and Akoya Biosciences.

Christopher Paustian reports no conflicts of interest.

Shu-Ching Chang reports no conflicts of interest.

Eloy Vetto reports no conflicts of interest.

Q. Vu reports no conflicts of interest.

V. Rajamanickam reports no conflicts of interest.

B. Bernard reports no conflicts of interest.

Y. Koguchi reports research funding from Bristol Myers Squibb, GlaxoSmithKline, and Shimadzu corporation outside the submitted work.

W. Redmond reports research support from: Bristol-Myers Squibb, Inhibrx, Veana Therapeutics,

Shimadzu, Galecto, and CanWell Pharma. Patents/Licensing fees: Galectin Therapeutics. Scientific

Advisory Boards: Vesselon, Medicenna, Veana Therapeutics.

B. A. Fox reports grants from Bristol Myers Squibb, Akoya Biosciences, Merck, and Incyte during the conduct of the study, personal fees from UbiVac, Turnstone, and Pfizer, Calidi, and PrimeVax outside the submitted work.

## Authors’ contributions

R. E. Sanborn: Clinical trial conception and design, funding acquisition, data curation, acquisition, and analysis, writing–original draft, edits, final draft, project administration. S.M. Jensen: Conceptualization, Data curation, formal analysis, validation, investigation, visualization, methodology, writing–review and editing C.C. Paustian: Methodology, formal analysis, writing–review and editing. S-C. Chang: formal analysis, Statistical analysis, Eloy Vetto: data curation and analysis. Q. Vu: Data analysis and formal analysis. V. Rajamanickam: Data curation, formal analysis, validation, investigation. B. Bernard: Formal analysis, supervision, Y. Koguchi: Formal analysis, supervision, visualization, writing–review and editing. W. Redmond: Formal analysis, supervision, writing–review and editing. B.A.Fox, Conceptualization, resources, supervision, funding acquisition, visualization, writing–review and editing

## Acknowledgements

Research support from Murdock Trust, Akoya Bioscience, Steve and Cindy Harder, Nancy Lematta, Lynn Loacker, the Chiles Foundation, and Providence Portland Medical Foundation. Note excellent assistance from Tyler Hulett, PhD and Gabriel Roman, PhD at CDI Labs in performing and assistance analyzing the PhIP-Seq data.

