## Supplemental Table 1 for "Provides Apparent Predictive Biomarker in a Phase I/II Study of Pembrolizumab With Gemcitabine in Patients with Previously-Treated Advanced Non-Small Cell Lung Cancer (NSCLC)"

Supplemental Table 1. Baseline Patient Characteristics

| **Median Age (Range)--years** | 64.5 (53-75) |
| --- | --- |
| **Gender Male/Female (%)** | 8 (50)/8 (50) |
| **Race/Ethnicity (%)** |  |
| Caucasian/Non-Hispanic | 14 (87.5) |
| Native Hawaiian/Other | 1 (6.25) |
| American or Alaskan Indian | 1 (6.25) |
| **Tumor Histology (%)** |  |
| Adenocarcinoma | 12 (75) |
| Squamous Cell Carcinoma | 1 (6.25) |
| NSCLC NOS | 3 (18.75) |
| **ECOG 0/1 (%)** | 2 (12.5)/14 (87.5) |
| **Tobacco use:** Never/Former/Current (%) | 3 (19)/9 (56)/4 (25) |
| **Prior Therapies** |  |
| **Radiation (%)** | 11 (68.75) |
| **Lines of Systemic Therapy 1/2/3 (%)** | 12 (75)/ 2 (12.5)/ 2 (12.5) |

Key: NOS: Not otherwise specified; ECOG: Eastern Cooperative Oncology Group
