## Supplemental Table 2 for "Provides Apparent Predictive Biomarker in a Phase I/II Study of Pembrolizumab With Gemcitabine in Patients with Previously-Treated Advanced Non-Small Cell Lung Cancer (NSCLC)"

Supplemental Table 2. Adverse Events attributed at least possibly related to study drugs in first 2 cycles

| **AE Grade** | **1/2** | **3** | **4** |
| --- | --- | --- | --- |
| **Hematologic:** |  |  |  |
| WBC Decreased | 1 | 2 |  |
| ANC Decreased | 3 | 3 |  |
| Lymphopenia |  | 1 |  |
| Anemia | 3 | 2 |  |
| Thrombocytopenia | 2 |  |  |
| **Non-Hematologic:** |  |  |  |
| Fatigue | 9 |  |  |
| Nausea | 9 |  |  |
| Flu like symptoms | 6 |  |  |
| Fever | 5 |  |  |
| Myalgia | 4 |  |  |
| Pain | 3 |  |  |
| Chills | 2 |  |  |
| Pneumonia | 2 |  |  |
| Edema | 2 |  |  |
| Cough | 1 |  |  |
| Vomiting | 1 |  |  |
| Infusion reaction | 1 |  |  |
| Arthralgia | 1 |  |  |
| Oral pain | 1 |  |  |
| Oral lesions | 1 |  |  |
| Upper respiratory infection |  | 1 |  |
| Wheezing | 1 |  |  |
| Hypotension | 1 |  |  |
| Dyspnea |  | 1 |  |
| Hypoxia |  | 1 |  |
| Hypoxic respiratory failure |  |  | 1 |
| Alveolar hemorrhage | 1 |  |  |
| Dry skin | 1 |  |  |
| Rash | 1 |  |  |
| Pruritis | 1 |  |  |
| Anorexia | 1 |  |  |
| Hyponatremia | 1 | 1 |  |
| Hyperglycemia | 1 |  |  |
| Hypomagnesemia | 1 |  |  |
| Alanine aminotransferase increased | 1 |  |  |
| Aspartate aminotransferase increased | 1 |  |  |
| Hematuria | 1 |  |  |
