## Supplemental Table 3 for "Provides Apparent Predictive Biomarker in a Phase I/II Study of Pembrolizumab With Gemcitabine in Patients with Previously-Treated Advanced Non-Small Cell Lung Cancer (NSCLC)"

Supplementary Table 3. Adverse Events attributed at least possibly to study drugs in subsequent cycles

| **AE Grade** | **1/2** | **3** |
| --- | --- | --- |
| **Hematologic:** |  |  |
| WBC Decreased |  | 1 |
| ANC Decreased | 2 | 2 |
| **Non-Hematologic:** |  |  |
| Pain | 3 |  |
| Myalgia | 3 |  |
| Fatigue | 2 |  |
| Arthralgia | 2 |  |
| Increased creatinine | 2 |  |
| Rash | 2 |  |
| Diarrhea | 1 |  |
| Pruritis | 2 |  |
| Edema | 2 |  |
| Hypophosphatemia | 2 |  |
| Constipation | 1 |  |
| Nausea | 1 |  |
| Vomiting | 1 |  |
| Sepsis |  | 1 |
| Pneumonia | 1 |  |
| Dyspnea | 1 | 1 |
| Neuropathy | 1 |  |
| Chest pain | 1 |  |
| Chest tightness | 1 |  |
| Muscle Spasms | 1 |  |
| Headache | 1 |  |
| Dry Skin | 1 |  |
| Sensitive Skin | 1 |  |
| Hyperpigmentation | 1 |  |
| Dry mouth | 1 |  |
| Dry eyes | 1 |  |
| Anosmia | 1 |  |
| Hypotension | 1 |  |
| Alopecia | 1 |  |
| Hematuria | 1 |  |
| Bruising | 1 |  |
