## Supplemental Figure 2 for "Provides Apparent Predictive Biomarker in a Phase I/II Study of Pembrolizumab With Gemcitabine in Patients with Previously-Treated Advanced Non-Small Cell Lung Cancer (NSCLC)"

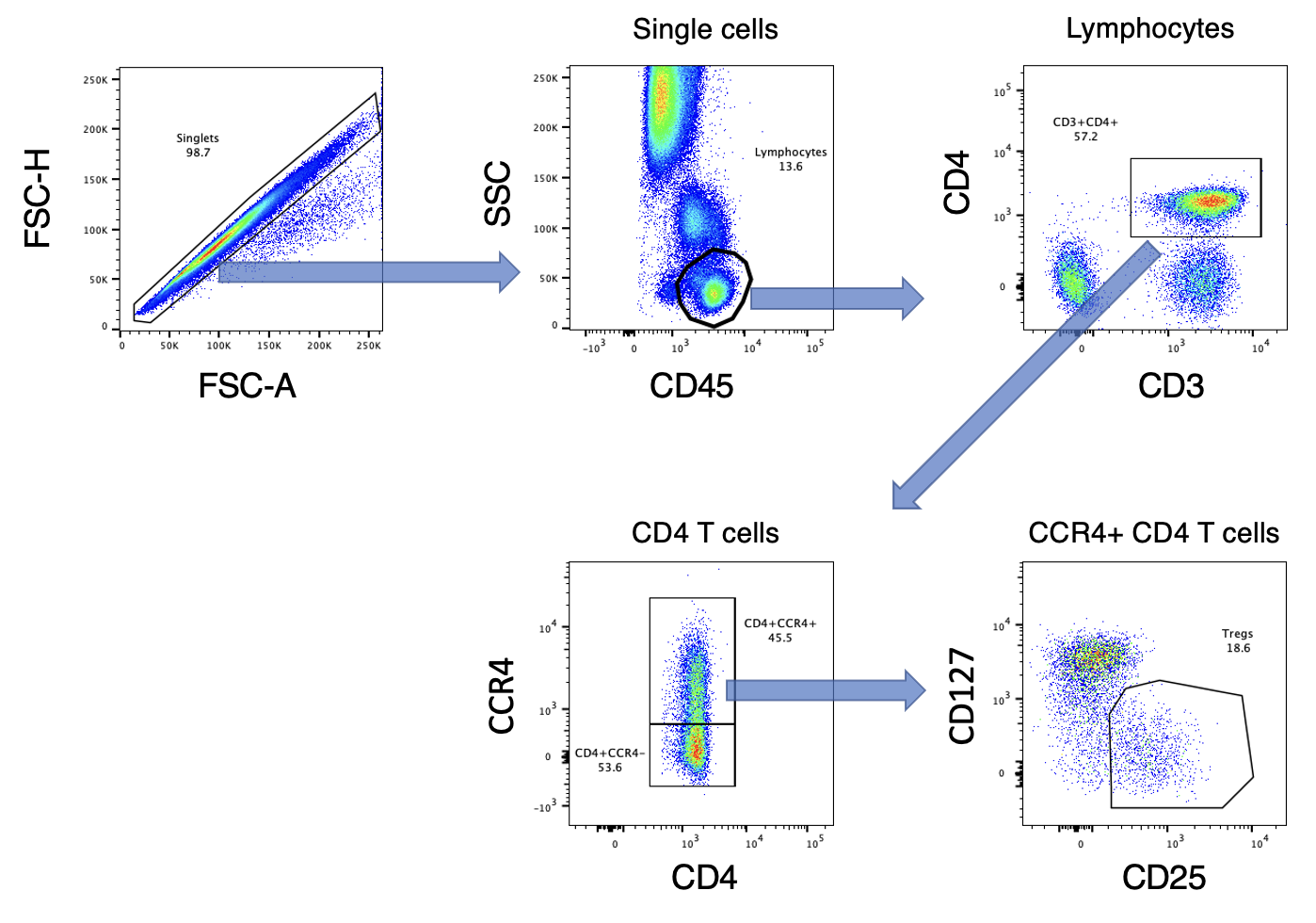

Supplemental Figure 2: Gating Strategy for Identifying Tregs. Tregs are defined as CD45+ CD3+ CD4+ CCR4 high CD127 low CD25 high cells within the lymphocyte gate. Arrows indicate which gated events were carried over to the next Flow-Cytometry plot.
