## Supplemental Figure 3 for "Provides Apparent Predictive Biomarker in a Phase I/II Study of Pembrolizumab With Gemcitabine in Patients with Previously-Treated Advanced Non-Small Cell Lung Cancer (NSCLC)"

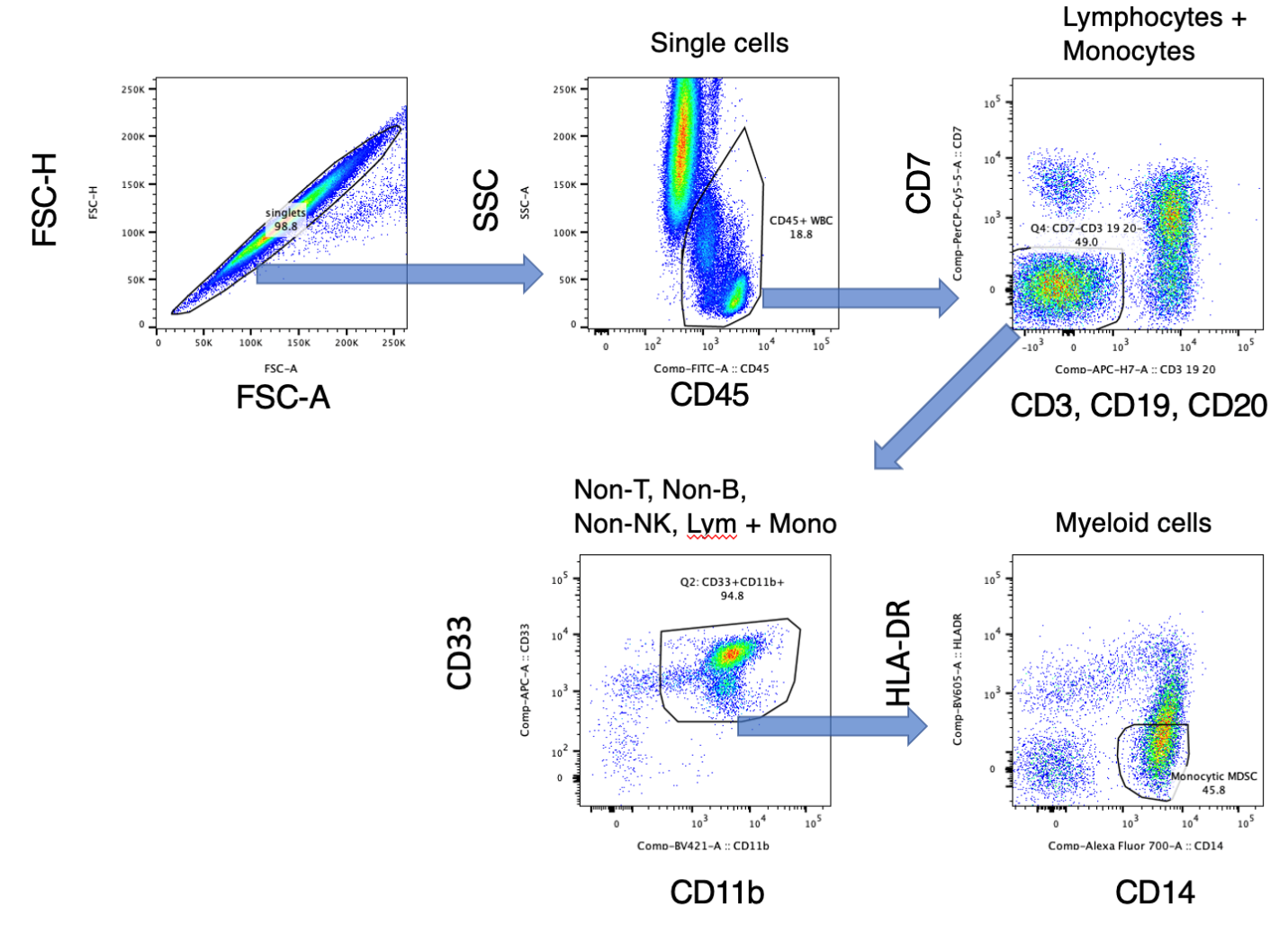

Supplemental Figure 3: Flow-cytometry gating of Monocytic MDSCs (Mo-MDSCs). Mo-MDSCs were defined as CD3- CD19- CD20- CD7- CD33+ CD11b+ CD14+ HLA-DR- within the lymphocytes + monocytes gate. Arrows indicate which gated events were carried over to the next Flow-Cytometry plot.
