## Supplemental Figure 4 for "Provides Apparent Predictive Biomarker in a Phase I/II Study of Pembrolizumab With Gemcitabine in Patients with Previously-Treated Advanced Non-Small Cell Lung Cancer (NSCLC)"

Supplemental Figure 4: CONSORT diagram

Screened: 22 Patients

Screen Fail: 6 Patients

Enrolled to Phase I: 6 Patients

Enrolled to Phase II: 10 Patients

Phase II Discontinuations: Disease Progression: 7 Patients Toxicity: 2 Patients Other: 1 Patient (Clinical decline with disease progression)

Phase I Discontinuations: Disease Progression: 4 Patients Toxicity: 1 Patient Pneumonitis (not DLT) Other: 1 Patient (Infection, clinical decline)
