## Supplemental Figure 5 for "Provides Apparent Predictive Biomarker in a Phase I/II Study of Pembrolizumab With Gemcitabine in Patients with Previously-Treated Advanced Non-Small Cell Lung Cancer (NSCLC)"

Supplemental Figure 5. Spider plot of response. % Change from baseline tumor measurement evaluated at follow-up timepoints. Each patient was assigned a color as indicated in the Legend to the right.

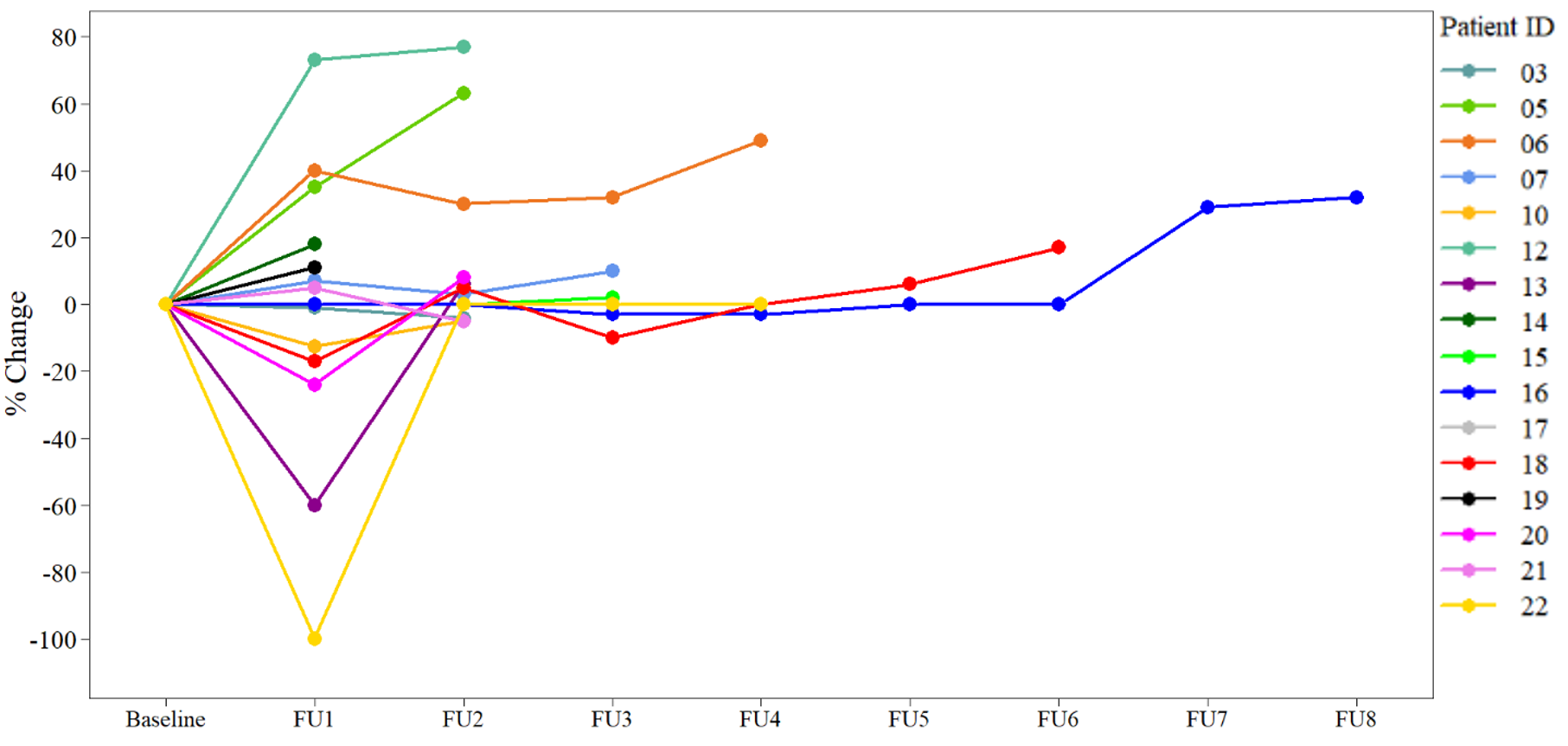

Supplemental Figure 5. Spider plot of response. % Change from baseline tumor measurement evaluated at follow-up timepoints. Each patient was assigned a color as indicated in the Legend to the right.
