## Supplemental Figure 6 for "Provides Apparent Predictive Biomarker in a Phase I/II Study of Pembrolizumab With Gemcitabine in Patients with Previously-Treated Advanced Non-Small Cell Lung Cancer (NSCLC)"

Supplemental Figure 6. Survival Based on PD-L1 Expression Levels. The Y-axis shows the percent of pre-treatment biopsies evaluated to be PD-L1 positive while the X-axis shows survival time in months. Patients progressing are indicated by (●), while those dying are marked by (x). Patients alive at the end of trial/follow-up are indicated with (○).

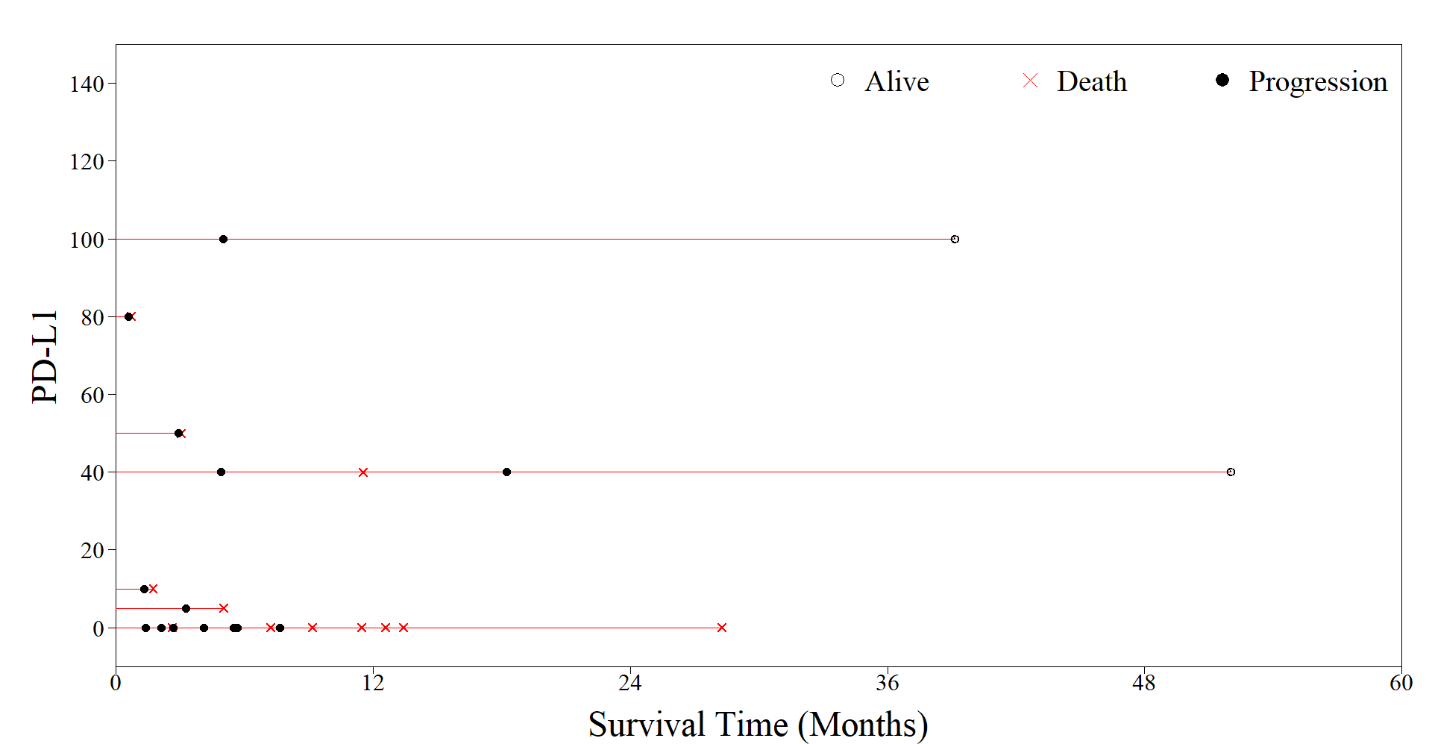
