## Supplemental Figure 7 for "Provides Apparent Predictive Biomarker in a Phase I/II Study of Pembrolizumab With Gemcitabine in Patients with Previously-Treated Advanced Non-Small Cell Lung Cancer (NSCLC)"

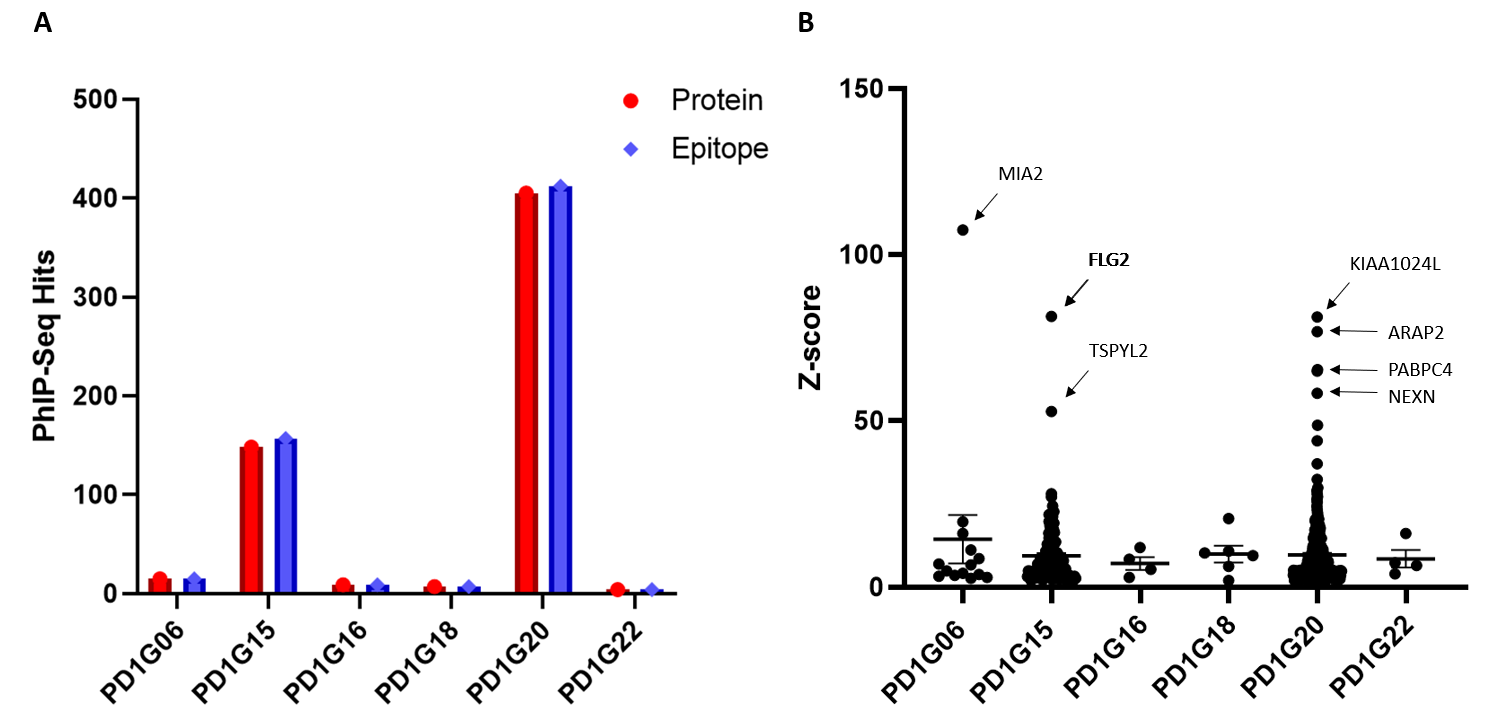

**Supplemental Figure 7.** Longitudinal data – Ab responses that were not detected at baseline. A) The number of antibody responses to distinct self-antigens (in red) or epitopes within self-antigens (blue) shown. B) Z-scores greater than 1.96 (95% confidence level) conveying the magnitude of hits to individual proteins compared to their beads only controls shown (Filled circles). Line and error bars represent mean and SEM respectively.
