## Supplemental Figure 9 for "Provides Apparent Predictive Biomarker in a Phase I/II Study of Pembrolizumab With Gemcitabine in Patients with Previously-Treated Advanced Non-Small Cell Lung Cancer (NSCLC)"

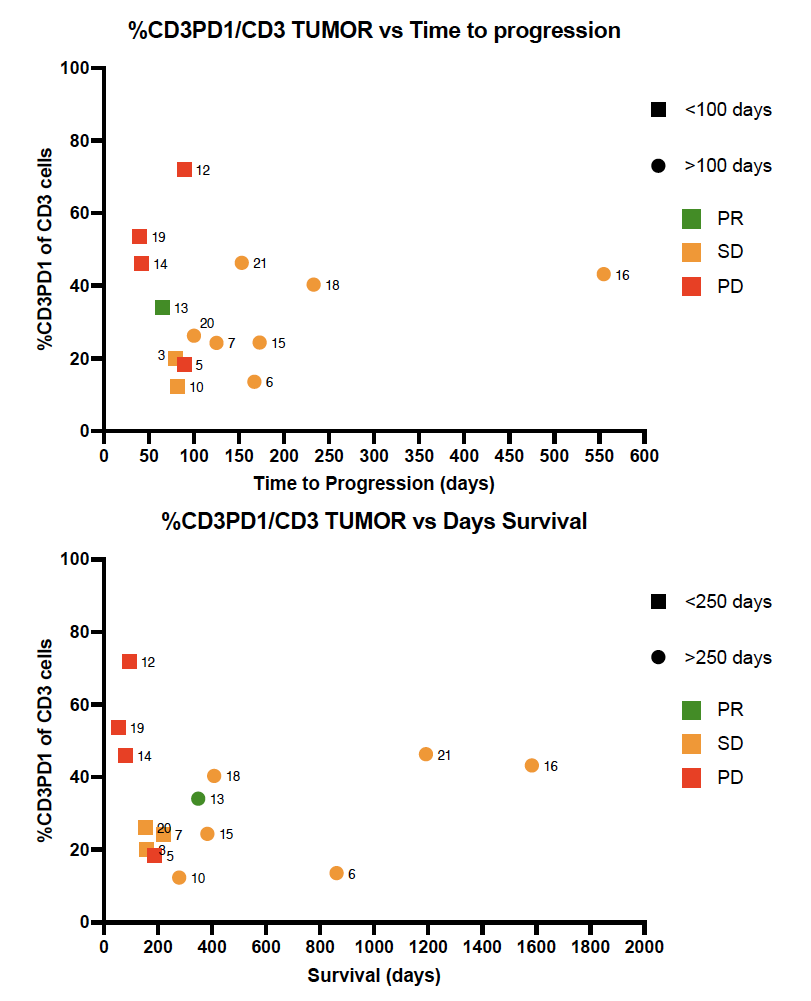

**Supplemental Figure 9**. *The percent of T cells (CD3^+^) expressing PD-1 plotted for the time to progression for each patient (upper figure) or plotted for patient survival (lower figure). green fill=partial response; orange fill=stable disease; red fill=progressive disease*
