## Supplemental Figure 10 for "Provides Apparent Predictive Biomarker in a Phase I/II Study of Pembrolizumab With Gemcitabine in Patients with Previously-Treated Advanced Non-Small Cell Lung Cancer (NSCLC)"

Supplemental Figure 10 . Study Schema

Phase I (6-36 patients), dose de-escalation

Pembrolizumab 200 mg IV every 3 weeks, for 2 years, or until disease progression

Gemcitabine 1250 mg/m^2^ IV Days 1, 8 every 3 weeks, for maximum 6 cycles

Phase II (10 patients)

Pembrolizumab MTD IV every 3 weeks, for 2 years, or until disease progression

Gemcitabine 1250 mg/m^2^ IV Days 1, 8 every 3 weeks, for maximum 6 cycles
