## Supplemental Figure 1 for "Provides Apparent Predictive Biomarker in a Phase I/II Study of Pembrolizumab With Gemcitabine in Patients with Previously-Treated Advanced Non-Small Cell Lung Cancer (NSCLC)"

**Support Provided By: Merck**

**Molecular Correlatives:**

Bernard A. Fox, Ph D.

Endowed Chair and Chief,

Molecular and Tumor Immunology

Earle A. Chiles Research Institute

Providence Portland Medical Center

4805 NE Glisan St, 2N35

Portland OR 97213

503-215-6841

### 1.0 TRIAL SUMMARY

| | |
|---|---|
| Abbreviated Title | **Phase I/II Study of Pembrolizumab With Gemcitabine in Patients with Previously-Treated Advanced Non-small Cell Lung Cancer (NSCLC)** |
| Trial Phase | I-II |
| Clinical Indication | Previously-treated advanced NSCLC |
| Trial Type | Open-label, non-randomized phase I study, followed by open-label non-randomized phase II |
| Type of control | Historical control |
| Route of administration | IV |
| Trial Blinding | N/A |
| Treatment Groups | Single arm, non-randomized, Pembrolizumab in combination with gemcitabine. |
| Number of trial subjects | 16-46 |
| Estimated duration of trial | 1 Year |
| Duration of Participation | 2 Years |

### 2.0 TRIAL DESIGN

#### 2.1 Trial Design

Phase I dose de-escalation component, followed by phase II nonrandomized study, compared with historical controls

##### 2.2 Trial Diagram

Phase I (6-36 patients), dose de-escalation

Pembrolizumab 200 mg iv every 3 weeks, for 2 years, or until disease progression

Gemcitabine 1250 mg/$m^2$ iv Days 1, 8 every 3 weeks, for maximum 6 cycles

↓

Phase II (10 patients)

Pembrolizumab MTD iv every 3 weeks, for 2 years, or until disease progression

Gemcitabine 1250 mg/$m^2$ iv Days 1, 8 every 3 weeks, for maximum 6 cycles

#### 3.0 OBJECTIVE(S) & HYPOTHESIS

##### 3.1 Primary Objective(s) & Hypothesis

(1) **Objective:** Safety

**Hypothesis:** The combination of pembrolizumab with gemcitabine will be safe and feasible in patients with previously-treated advanced NSCLC.

##### 3.2 Secondary Objective(s) & Hypothesis

(1) **Objective**: Progression-free survival

**Hypothesis**: The combination of pembrolizumab with gemcitabine will improve progression-free survival over that seen with either gemcitabine or pembrolizumab administered as single-agent therapy for patients with previously-treated advanced NSCLC, in comparison with historical controls.

(2) Objective: Overall survival

(3) Objective: Response rate

##### 3.3 Exploratory Objective

(1) **Objective:** Correlative analysis of outcomes retrospectively compared with tumor PD-L1 expression, tumor immune score, and immune response as assessed by serial protein arrays.

#### 4.0 BACKGROUND & RATIONALE

##### 4.1 Background

Non-small cell lung cancer (NSCLC) is the most common cancer killer worldwide, and in the United States alone in 2014, an estimated 159,000 people will die from lung cancer (1). The majority of patients with NSCLC are diagnosed at an advanced stage, when goals of therapy are palliative, but not curative. Palliative chemotherapy has been shown to help prolong life and improve quality of life for patients with good performance status with advanced disease, but duration of disease control with chemotherapy is still short, and average survival with palliative chemotherapy is still only approximately 12 months (2, 3).

Gemcitabine is an active agent for patients with NSCLC, commonly used in the first-line setting for advanced squamous cell lung cancer in combination with platinum drugs, or as single agent therapy in the salvage setting for patients with any histology of NSCLC. Gemcitabine is used frequently for elderly patients with NSCLC due to its relatively lower toxicity profile. As it does not cause neuropathy, it is an attractive agent for patients with either preexisting neuropathy from other medical conditions, or chemotherapy-induced neuropathy.

Single agent gemcitabine has been shown to have progression-free survival (PFS) ranging between 2.5-4.25 months, and median survival of 4.4-7 months in the first-line setting for patients over the age of 70 (4-6). Phase III trials of gemcitabine in the second-line setting for NSCLC have not been performed, but in phase II studies median time to progression is 2.5 months, with median overall survival 9.5 months, and overall response rate 18.5% (7).

Chemotherapy agents alone have reached a plateau of therapeutic efficacy. Targeted therapy agents have made a significant impact in survival and quality of life for patients with advanced NSCLC, particularly for those patients with sensitizing mutations (8, 9), however, resistance ultimately develops and patients inevitably experience disease progression on targeted therapies. There is a significant need for agents able to induce long-term responses and survival, with manageable side effects.

##### 4.1.1 Pharmaceutical and Therapeutic Background

Refer to the Investigator's Brochure (IB) for detailed background information on pembrolizumab.

The importance of intact immune surveillance in controlling outgrowth of neoplastic transformation has been known for decades [10]. Accumulating evidence shows a correlation between tumor-infiltrating lymphocytes (TILs) in cancer tissue and favorable prognosis in various malignancies [11-15]. In particular, the presence of CD8+ T-cells and the ratio of CD8+ effector T-cells / FoxP3+ regulatory T-cells seems to correlate with improved prognosis and long-term survival in many solid tumors.

The PD-1 receptor-ligand interaction is a major pathway hijacked by tumors to suppress immune control. The normal function of PD-1, expressed on the cell surface of activated T-cells under healthy conditions, is to down-modulate unwanted or excessive immune responses, including autoimmune reactions. PD-1 (encoded by the gene Pdcd1) is an Ig superfamily member related to CD28 and CTLA-4 which has been shown to negatively regulate antigen receptor signaling upon engagement of its ligands (PD-L1 and/or PD-L2) [16; 17]. The structure of murine PD-1 has been resolved [18]. PD-1 and family members are type I transmembrane glycoproteins containing an Ig Variable-type (V-type) domain responsible for ligand binding and a cytoplasmic tail which is responsible for the binding of signaling molecules. The cytoplasmic tail of PD-1 contains 2 tyrosine-based signaling motifs, an immunoreceptor tyrosine-based inhibition motif (ITIM) and an immunoreceptor tyrosine-based switch motif (ITSM). Following T-cell stimulation, PD-1 recruits the tyrosine phosphatases SHP-1 and SHP-2 to the ITSM motif within its cytoplasmic tail, leading to the dephosphorylation of effector molecules such as CD3ζ, PKCθ and ZAP70 which are involved in the CD3 T-cell signaling cascade [16, 19-21]. The mechanism by which PD-1 down modulates T-cell responses is similar to, but distinct from that of CTLA-4 as both molecules regulate an overlapping set of signaling proteins [22; 23]. PD-1 was shown to be expressed on activated lymphocytes including peripheral CD4+ and CD8+ T-cells, B-cells, T regs and Natural Killer cells [24; 25]. Expression has also been shown during thymic development on CD4-CD8- (double negative) T-cells as well as subsets of macrophages and dendritic cells [26]. The ligands for PD-1 (PD-L1 and PD-L2) are constitutively expressed or can be induced in a variety of cell types, including non-hematopoietic tissues as well as in various tumors [22, 27-29]. Both ligands are type I transmembrane receptors containing both IgV- and IgC-like domains in the extracellular region and contain short cytoplasmic regions with no known signaling motifs. Binding of either PD-1 ligand to PD-1 inhibits T-cell activation triggered through the T-cell receptor. PD-L1 is expressed at low levels on various non-hematopoietic tissues, most notably on vascular endothelium, whereas PD-L2 protein is only detectably expressed on antigen-presenting cells found in lymphoid tissue or chronic inflammatory environments. PD-L2 is thought to control immune T-cell activation in lymphoid organs, whereas PD-L1 serves to dampen unwarranted T-cell function in peripheral tissues [22]. Although healthy organs express little (if any) PD-L1, a variety of cancers were demonstrated to express abundant levels of this T-cell inhibitor. PD-1 has been suggested to regulate tumor-specific T-cell expansion in subjects with melanoma (MEL) [30]. This suggests that the PD-

1/PD-L1 pathway plays a critical role in tumor immune evasion and should be considered as an attractive target for therapeutic intervention.

Pembrolizumab (previously known as MK-3475; SCH 900475) is a potent and highly selective humanized monoclonal antibody (mAb) of the IgG4/kappa isotype designed to directly block the interaction between PD-1 and its ligands, PD-L1 and PD-L2 without ADCC or CDC activity.

###### 4.1.2 Preclinical and Clinical Trial Data

Refer to the Investigator's Brochure for Preclinical and Clinical data.

##### 4.2 Rationale

###### 4.2.1 Rationale for the Trial and Selected Subject Population

In a large phase I study involving 296 patients, anti-PD1 (Nivolumab) demonstrated responses in patients with melanoma, renal cell carcinoma, and NSCLC. Patients received a dose range of anti-PD1 of 1.0-10.0 mg/kg intravenously every two weeks (of 8-week treatment cycles) for up to two years (12 cycles). For the 76 patients with heavily-pretreated advanced NSCLC (55% having received at least 3 lines of prior therapy), the response rate was 18%. Progression-free survival in this population was an impressive 26 weeks, with 7% of patients experiencing prolonged stable disease (31). In long-term follow up, the estimated duration of response was 74 weeks, with ongoing response in 55% of the responding patients. 1-year overall survival was 42%, with 2-year overall survival 14% (32).

A maximum tolerated dose of Nivolumab was not reached, with 5% of patients discontinuing therapy due to treatment-related adverse events. Toxicity did not appear to be dose-related. For the overall population, the incidence of grade 3 or 4 treatment-related adverse events was 14%. Serious adverse events attributed to drug occurred in 11% of patients. Adverse events which were potentially immune related included vitiligo, colitis, hepatitis, hypophysitis, thyroiditis, and pneumonitis, with grade 3 or 4 toxicity occurring in 6% of patients (18 patients total). Of the grade 3 or 4 adverse events, three patients (1%) developed pneumonitis, three developed colitis (1%), two patients each developed ALT or AST elevations (1% each). Pruritis, macular rash, elevated thyroid-stimulating hormone, hypo- and hyperthyroidism were recorded in 1 patient each. One patient developed a grade 3 or 4 infusion-related reaction. For patients developing pneumonitis, there was no relationship seen between dose level or number of doses administered, or to tumor type. Ultimately, three drug-related pneumonitis deaths were reported, with two of these occurring in patients with NSCLC. Patients with mild or moderate pneumonitis were treated with glucocorticoids, although patients with more severe pneumonitis required more aggressive immune suppression, with infliximab or mycophenolate (31). The investigators concluded that although pneumonitis was observed, overall the toxicities with the study drug were acceptable, and the drug was safe to be administered in the outpatient setting. The unprecedented results of an immunotherapy approach in NSCLC, along with other tumors, has prompted widespread investigation and development of immunotherapy agents, including anti-PDL1 and anti-PD1 agents (www.clinicaltrials.gov).

Pembrolizumab is being investigated in a number of different therapeutic settings currently (www.clinicaltrials.gov). The KEYNOTE-001 phase I study in previously-treated advanced NSCLC evaluated pembrolizumab at 10 mg/kg iv every 2 or every 3 weeks, and evaluated patients for response based upon tumor PD-L1 expression. PFS was not significantly different between patients with PD-L1 positive or negative tumors, with median PFS 10 weeks for PD-L1 negative tumors and 11 weeks for PD-L1 positive tumors based upon independent central review using RECIST v1.1 (16 weeks for both based upon investigator review using irRC; 3). Overall response rate was 21% for patients with PD-L1 positive tumors treated with every 3 week dosing, with duration of response of 31 weeks based upon independent central review (33).

Although gemcitabine as a chemotherapy drug has been generally considered to be purely immunosuppressive, preclinical studies have indicated that gemcitabine increases antigen cross-presentation, as well as increases the percentage of intratumoral primed tumor-specific CD8 T cells, without increasing functional tolerance (34). Gemcitabine combined with nonspecific immunotherapy (anti-CD40 antibody) in subsequent preclinical experiments using a mesothelioma murine model demonstrated synergism between the two agents. It was postulated that gemcitabine induced immunological priming via increasing tumor antigen cross-presentation as well as upregulating antigen-presenting cells through induction of apoptosis, thereby improving subsequent antitumor immune response induced by immunotherapy (35).

We thus propose a feasibility study of pembrolizumab in combination with gemcitabine for patients with previously-treated advanced NSCLC. Should pembrolizumab demonstrate safety and tolerability in this patient population, as well as efficacy signal, this would support further investigation with a randomized trial with the potential for establishing a new standard of care for a very large patient population with significant need.

##### 4.2.2 Rationale for Dose Selection/Regimen/Modification

An open-label Phase I trial (Protocol 001) is being conducted to evaluate the safety and clinical activity of single agent pembrolizumab. The dose escalation portion of this trial evaluated three dose levels, 1 mg/kg, 3 mg/kg, and 10 mg/kg, administered every 2 weeks (Q2W) in subjects with advanced solid tumors. All three dose levels were well tolerated and no dose-limiting toxicities were observed. This first in human study of pembrolizumab showed evidence of target engagement and objective evidence of tumor size reduction at all dose levels (1 mg/kg, 3 mg/kg and 10 mg/kg Q2W). No MTD has been identified to date. Recent data from other clinical studies within the pembrolizumab program has shown that a lower dose of pembrolizumab and a less frequent schedule may be sufficient for target engagement and clinical activity.

PK data analysis of pembrolizumab administered Q2W and Q3W showed slow systemic clearance, limited volume of distribution, and a long half-life (refer to IB). Pharmacodynamic

data (IL-2 release assay) suggested that peripheral target engagement is durable (>21 days). This early PK and pharmacodynamic data provides scientific rationale for testing a Q3W dosing schedule.
Available pharmacokinetic results in subjects with melanoma, NSCLC, and other solid tumor types support a lack of meaningful difference in pharmacokinetic exposures obtained at a given dose among tumor types. Population PK analysis has been performed and has confirmed the expectation that intrinsic factors do not affect exposure to pembrolizumab to a clinically meaningful extent. Taken together, these data support the use of lower doses (with similar exposure to 2 mg/kg Q3W) in all solid tumor indications. 2 mg/kg Q3W is being evaluated in NSCLC in PN001, Cohort F30 and PN010, and 200 mg Q3W is being evaluated in head and neck cancer in PN012, which are expected to provide additional data supporting the dose selection.

Selection of 200 mg as the appropriate dose for a switch to fixed dosing is based on simulation results indicating that 200 mg will provide exposures that are reasonably consistent with those obtained with 2 mg/kg dose and importantly will maintain individual patient exposures within the exposure range established in melanoma as associated with maximal clinical response. A population PK model, which characterized the influence of body weight and other patient covariates on exposure, has been developed using available data from 476 subjects from PN001. The distribution of exposures from the 200 mg fixed dose are predicted to considerably overlap those obtained with the 2 mg/kg dose, with some tendency for individual values to range slightly higher with the 200 mg fixed dose. The slight increase in PK variability predicted for the fixed dose relative to weight-based dosing is not expected to be clinically important given that the range of individual exposures is well contained within the range of exposures shown in the melanoma studies of 2 and 10 mg/kg to provide similar efficacy and safety. The population PK evaluation revealed that there was no significant impact of tumor burden on exposure. In addition, exposure was similar between the NSCLC and melanoma indications. Therefore, there are no anticipated changes in exposure between different tumor types and indication settings.

Table 1. Dose Schema for Pembrolizumab and Gemcitabine

| 21-day Cycles | Cycle 1/Day 1 | C1/D8 | C2/D1 | C2/D8 |
|---|---|---|---|---|
| Study Drug | | | | |
| Pembrolizumab[a] 200 mg intravenously | X | | X | |
| Gemcitabine[b] 1250 $mg/m^2$ intravenously | X | X | X | X |

a. Pembrolizumab will be administered on Day 1 every 21 days (3 weeks) for 2 years, or until confirmed disease progression or prohibitive toxicity, whichever comes first.

b. Gemcitabine will be administered on Days 1 and 8 of each 21 day (3 week) cycle for a maximum of 6 cycles, or until disease progression or prohibitive toxicity, whichever comes first.

Table 2. Dose modifications due to toxicity.

| | |
|---|---|
| **Pembrolizumab** | |
| Initial dose | 200 mg Day 1 every 21 days |
| -1 Dose level | 100 mg Day 1 every 21 days |
| **Gemcitabine** | |
| Initial dose | 1250 $mg/m^2$ Days 1, 8 every 21 days |
| -1 Dose level | 1000 $mg/m^2$ Days 1, 8 every 21 days |
| -2 Dose level | 800 $mg/m^2$ Days 1, 8 every 21 days |

##### 4.2.3 Rationale for Endpoints

Pembrolizumab has not yet been evaluated in combination with gemcitabine in patients with previously-treated advanced NSCLC. We propose that the combination will be safe and feasible. The primary objective of this study will be to establish safety of the combination in this patient population.

###### 4.2.3.1 Efficacy Endpoints

We propose that pembrolizumab in combination with gemcitabine will improve PFS over gemcitabine alone for patients with advanced NSCLC. We propose that the combination of pembrolizumab and gemcitabine will be synergistic, with potential based upon the phase II portion of this study for further evaluation with a larger randomized trial in this population with significant unmet need.

We further propose that the combination of pembrolizumab and gemcitabine will improve overall survival for patients with previously-treated advanced NSCLC compared with gemcitabine alone, with potential based upon the phase II portion of this study for further evaluation with a larger randomized trial in this population with significant unmet need.

We propose that the overall response rate with the combination of pembrolizumab and gemcitabine will be improved compared with gemcitabine alone for patients with previously-treated advanced NSCLC.

###### 4.2.3.2 Biomarker Research

**Immunoscore and immunoprofiling evaluation:**

We hypothesize that checkpoint blockade with anti-PD-1 will only be effective in patients with a pre-existing anti-cancer immune response. We also hypothesize that the objective assessment of immune infiltrate present in the cancer of patients enrolled on this study will inform subsequent trial design, by identifying a predictive biomarker of response. In studies of patients with resected colorectal cancer, tumor infiltration by CD3+ and CD8+ cytotoxic T cells as quantified by immunohistochemical (IHC) staining, digital imaging and an objective assessment of the tumor center and invasive margins (the immunoscore) was shown to correlate with survival and tumor recurrence (36). High densities of CD8+ T cells at the center and invasive margins (high immunoscore) correlated with earlier T-stage tumors, while low density infiltration (low immunoscore) correlated with larger tumors and disease relapse. In a recent study, a positive immunoscore independently correlated with significantly lower risk of disease recurrence and improved survival and provided a significantly better prognostic correlation than TNM staging (37). An international consortium has begun a retrospective analysis of 5000 patients to validate the CD3+ CD8+ immune score as a prognostic biomarker in patients with colon cancer and Drs. Fox and Bifulco at our Institute serve on the Society for Immunotherapy of Cancer (SITC) Taskforce that is coordinating and standardizing staining for the 23 participating centers in 17 countries (38). Thus, our group is experienced in this assessment and well suited to perform the proposed studies (39).

In NSCLC, resected stage I adenocarcinomas were evaluated for infiltrating immune cells (helper, cytotoxic, and memory T cells, regulatory T cells, B cell, NK cell, and macrophage infiltration) in the tumor and surrounding microenvironment. A high density of regulatory T cells in the stroma (but not the tumor) was associated with higher risk of tumor recurrence, and an inverse correlation of regulatory T cells to CD3+ T cells in stroma correlated strongly with risk of recurrence (40).

Currently, there is not a definitive biomarker that predicts with absolute reliability benefit (or lack thereof) with immunotherapy agents, including pembrolizumab. Although tumor PD-L1 expression is under investigation as a likely biomarker, tumor responses have been demonstrated in patients treated with anti-PD-1 and anti-PD-L1 agents whose tumors are considered negative for PD-1 and PD-L1 expression. Thus, further investigation to develop an understanding of those patients potentially more (or less) likely to experience a benefit from immunotherapy, including pembrolizumab, is critical. Assessment of a pre-treatment predictive assessment of prognosis or potential benefit from immunotherapy, such as the immunoscore, carries a significant potential that could be translated into further investigations in other disease stages with pembrolizumab or other immunotherapy agents.

**Humoral Immune response assessment – 9000 Protein spotted Arrays:**

What does the immune system of a patient recognize if they experience a PR or CR following treatment with pembrolizumab? A major question in the field is whether these responses are targeting mutated or non-mutated proteins. Monitoring for immune response with serial protein arrays will allow for assessment of patients with and without a pre-existing humoral immune response (as demonstrated by changes in immune studies), and exploratory analyses comparing outcomes between those patients generating immune responses and those who do not. This may allow for development of further predictors of benefit with immunotherapy agents. Additionally, these studies may help determine the targets of immune destruction, aiding in assessing pembrolizumab's mechanism of action. Our group recently used these arrays to assess potential cancer antigens in a patient with advanced prostate cancer who experienced a durable CR (6 years+) following treatment with ipilimumab (41).

**Immunophenotyping of whole blood:**

Our Institute has invested substantial resources to develop an outstanding immunological monitoring laboratory under the direction of Dr. Keith Bahjat. The proposed studies will evaluate whole blood samples for changes in T cell subsets. The four panels evaluate: 1) T cell cocktail - CD4 and CD8 T cells for absolute numbers of naïve. memory, central memory and effector memory subsets, 2) T Help cocktail, 3) T reg cocktail and 4) a DCNK Cocktail. Together, these 4 panels detailed in the budget justification section, provide a solid overview of changes in these immune cell subsets. Additionally, blood from the pre and week 12 interval will be cryopreserved for possible functional studies.

### 5.0 METHODOLOGY

#### 5.1 Entry Criteria

##### 5.1.1 Diagnosis/Condition for Entry into the Trial

Women or men with advanced, histologically proven NSCLC. Mixed histologies are allowed as long as no small cell elements are seen. Advanced disease is defined as disease not treatable with curative intent, such as stage IV disease, or recurrent stage III disease.

##### 5.1.2 Subject Inclusion Criteria

1. Women or men with advanced, histologically proven NSCLC. Mixed histologies are allowed as long as no small cell elements are seen. Advanced disease is defined as disease not treatable with curative intent, such as stage IV disease, or recurrent stage III disease.

2. Patients must have received at least one but no more than three prior systemic therapies for advanced disease. Prior neoadjuvant or adjuvant chemotherapy will not be included in the assessment as a prior chemotherapeutic regimen, unless disease recurrence was less than 12 months after completing therapy.

3. Per Good Clinical Practice, any toxicity related to prior therapies that, in the opinion of the investigator, would potentially be worsened with anti-PD1 therapy or gemcitabine should be resolved to less than Grade 1.

**4.** Eastern Cooperative Oncology Group (ECOG) performance status 0 or 1.

**5.** Age ≥ 18 years.

**6.** Women of childbearing potential must have a negative pregnancy test and must avoid becoming pregnant while on treatment. Men must avoid fathering a child while on treatment.

**7.** Ability to give informed consent and comply with the protocol.

**8.** Anticipated survival minimum 3 months.

**9.** Prior therapy with investigational agents must have been completed at least 3 weeks prior to study enrollment.

**10.** Patients must have normal organ and marrow function as defined by:

| | |
|---|---|
| a. Leukocytes | ≥ 3,000/µL |
| b. Platelets | ≥ 100,000/µL |
| c. Total bilirubin | Within normal institutional limits |
| d. AST (SGOT)/ALT (SGPT) | ≤ 2 x institutional upper limit of normal |

11. Measurable disease by RECIST 1.1 criteria.

12. Treated brain metastases will be allowed, provided they are asymptomatic. Radiation treatment for brain metastases must have been completed at least 2 weeks prior to enrollment. Patients must demonstrate stable symptoms and seizure control on a consistent dose of anticonvulsants (if deemed necessary) for at least 2 weeks prior to enrollment. Patients must have been either off corticosteroids, or on a stable or decreasing dose of </= 10mg Prednisone or Prednisone equivalent daily for at least 2 weeks. Treatment for brain metastases may include whole brain radiotherapy (WBRT), radiosurgery (RS; Gamma Knife, KINAC, or equivalent), neurosurgical resection with radiation, or a combination as deemed appropriate by the treating physician.

13. Radiation for symptomatic lesions outside the CNS must have been completed at least 2 weeks prior to study enrollment. If measurable disease is within the radiation field, there must be evidence of clear progression (using RECIST criteria) at the time of study enrollment.

##### 5.1.3 Subject Exclusion Criteria

1. Prior therapy with any anti-PD-1, anti-PD-L1, or anti-CTLA4 antibody.

2. Prior therapy with gemcitabine.

3. Prior complications from radiation, such as history of radiation pneumonitis or pulmonary edema, that, in the opinion of the investigator, may have risk of increasing toxicity with anti-PD1 therapy.

4. History of (non-infectious) pneumonitis that required steroids, or current pneumonitis.

5. Evidence of interstitial lung disease.

6. Previous allergic reaction to gemcitabine.

7. Active autoimmune disease except vitiligo or stable hypothyroidism.

8. Active and ongoing steroid use greater than 10mg Prednisone or Prednisone equivalent daily. Non-systemically absorbed treatments (such as inhaled or topical steroid therapy for asthma, COPD, allergic rhinitis) are allowed.

**9.** Active other malignancy, except for controlled basal cell skin carcinoma.

**10.** HIV positive and/or Hepatitis B or C positive.

**11.** Other medical or psychiatric conditions that in the opinion of the Principal Investigator would preclude safe participation in this protocol.

#### 5.2 Trial Treatments

The treatment to be used in this trial is outlined below in Table 3.

Table 3 Trial Treatment—Phase I portion, initial dose cohort.

| Drug | Dose/Potency | Dose Frequency | Route of Administration | Regimen/Treatment Period | Use |
|---|---|---|---|---|---|
| Pembrolizu-mab | 200 mg | Q21 days | IV infusion | 24 Months | Experimental |
| Gemcitabine | 1250 mg/m$^2$ | Days 1, 8 of 21- | IV infusion | 6 cycles | Standard of Care |

| Drug | Dose/Potency | Dose Frequency | Route of Administration | Regimen/Treatment Period | Use |
|---|---|---|---|---|---|
| | | day schedule | | | |
| The pembrolizumab dosing interval may be increased due to toxicity as described in Section 5.2.1.2. | | | | | |

##### 5.2.1 Dose Selection/Modification

###### 5.2.1.1 Dose Selection

The rationale for selection of doses to be used in this trial is provided in Section 4.0 – Background and Rationale.

Details on the dose preparation and administration are provided in the Procedures Manual.

Dose Limiting Toxicity (DLT) will be defined as grade 4 hematologic, or grade 3 or 4 non-hematologic toxicity using NCI Common Terminology Criteria for Adverse Events v4.0 that in the opinion of the investigator is considered at least possibly related to treatment. Any grade 3 or 4 hematologic or non-hematologic toxicity will result in stopping of therapy until the toxicity has resolved to grade 1 or less. If treatment needs to be held for longer than 3 weeks from the scheduled dose due to toxicity, this is also considered DLT. If resolution of the toxicity to grade 2 or less has not occurred within four weeks, the patient will be withdrawn from study.

The initial phase I cohort will enroll a maximum of 6 patients.

If DLT felt to be attributed to pembrolizumab is observed in two or more patients, then a dose de-escalation will occur, with six further patients enrolled receiving pembrolizumab 100 mg iv every 3 weeks. If DLT felt to be attributed to gemcitabine is observed in two or more patients, then a dose de-escalation of gemcitabine will occur, with six further patients enrolled receiving the next lower dose of gemcitabine (See Dose Modifications, Table 2, Section 4.2.2). Dose de-escalation of either drug may occur separately from the other depending upon attribution of toxicity.

If DLT felt to be attributed to pembrolizumab is observed in two or more patients in the dose de-escalated cohort, the combination will be considered not feasible, and accrual will be halted.

After the first 2 patients are enrolled and start therapy, further enrollment will be held until both patients complete 6 weeks of therapy. If DLT is seen in one or fewer patients, then accrual will reopen. Enrollment will then be held again after enrollment of 2 more patients until both patients complete 6 weeks of therapy. If dose-de-escalation criteria are not met, then enrollment will open to complete the phase I cohort with the same dose of pembrolizumab to a maximum of 6 patients total.

If no DLTs are seen, then the phase II portion will commence enrollment.

The MTD in the phase I portion of the study will be used in the phase II portion of the trial.

###### 5.2.1.2 Dose Modification

Dose adjustments of gemcitabine should be made according to the greatest degree of toxicity as recorded according to the CTCAE Version 4.0, and are listed in Table 4. Dose re-escalation after dose reduction for toxicity of either drug will not be allowed. There will be no more than two dose reductions allowed for gemcitabine and no more than one dose reduction of pembrolizumab.

A maximum of a six-week treatment delay will be permitted to allow recovery from gemcitabine-induced toxicities. If a subject experiences recurring toxicities despite 2 dose reductions of gemcitabine, treatment should be discontinued. The subject will continue to be followed for resolution of toxicity as well as for disease progression and survival.

Subjects who require dose delays of gemcitabine for chemotherapy-related toxicity may continue to receive pembrolizumab on schedule, provided the toxicity is not thought to be due to pembrolizumab (See Table 5 for management of pembrolizumab-related toxicities).

Table 4. Gemcitabine dose modification guidelines for drug-related toxicities.

| **Hematologic Toxicities** | | |
|---|---|---|
| **Neutropenia: Day 1** | 1) Grade 1<br>2) Grade 2<br>3) Grade ≥3 | 1) No dose modification<br>2) Hold dose until ≤ Grade 1*<br>3) Hold dose until ≤ Grade 1*<br>Reduce dose -1 level* |
| **Neutropenia: Days 8** | 1) Grade 1<br>2) Grade 2<br>3) Grade ≥ 3 | 1) No dose modification<br>2) Hold dose until ≤ Grade 1*<br>3) Hold dose until ≤ Grade 1*<br>Reduce dose -1 level* |
| **Febrile Neutropenia**<br>(one reading of oral temp. >38.5°C, or three readings of oral temperature >38.0° C in a 24-hour period) | 1) First event<br>2) Second event<br>3) Third event | 1) Administer G-CSF and delay treatment cycle a minimum of 1 week. Hospitalization and antibiotic therapy as clinically indicated.<br>Discontinue treatment if ANC does not recover within 28 days.<br>Reduce dose -1 level* |

| | | |
|---|---|---|
| | | 2) Reduce dose –2 level*<br>3) Discontinue treatment |
| **Grade 3 or 4 Infection without Neutropenia** | 1) First event<br>2) Second event<br>3) Third event | 1) Consider prophylactic antibiotics with subsequent cycles<br>2) Reduce to –1 dose level*<br>3) Reduce to –2 dose level* |
| **Anemia** | 1) Grade 1<br>2) Grade ≥2 | 1) No dose reduction<br>2) Transfuse red blood cell products and administer erythropoietin products at investigator discretion |
| **Thrombocytopenia** | 1) ≥100,000/µL<br>2) ≤ 100,000/µL | 1) No dose reduction<br>2) Hold gemcitabine until platelet count recovers to ≥100,000/µL (maximum 28 days);*Reduce dose –1 level* |
| Non-Hematologic Toxicities | | |
| **Grade 1** | N/A | No change in therapy |
| **Grade 2** | N/A | Hold gemcitabine until ≤ Grade 1 |
| **Grade 3-4** | 1. First event<br>2. Second event | Supportive care as necessary<br>Hold gemcitabine until ≤ Grade 1<br>1. Reduce dose -1 level*<br>2. Reduce dose -2 level* |

* Doses held during a course of therapy will be deleted and will not be administered at a later time. If a dose reduction is required day 8, the dose reduction will be permanent. If treatment is held for over 4 weeks due to hematologic toxicity, the patient will be removed from the study.

Pembrolizumab will be withheld for drug-related Grade 3 or 4 hematologic toxicities, non-hematological toxicity ≥ Grade 3 including laboratory abnormalities, and severe or life-threatening AEs as per Table 5 below.

Table 5: Dose Modification and Toxicity Management Guidelines for Immune-related AEs Associated with Pembrolizumab

**General instructions:**

1. Corticosteroid taper should be initiated upon AE improving to Grade 1 or less and continue to taper over at least 4 weeks.
2. For situations where pembrolizumab has been withheld, pembrolizumab can be resumed after AE has been reduced to Grade 1 or 0 and corticosteroid has been tapered. Pembrolizumab should be permanently discontinued if AE does not resolve within 12 weeks of last dose or corticosteroids cannot be reduced to ≤10 mg prednisone or equivalent per day within 12 weeks.
3. For severe and life-threatening immune-related AEs (irAEs), IV corticosteroid should be initiated first followed by oral steroid. Other immunosuppressive treatment should be initiated if irAEs cannot be controlled by corticosteroids.

| **Immune-related AEs** | **Toxicity grade or conditions (CTCAEv4.0)** | **Action taken to pembrolizumab** | **irAE management with corticosteroid and/or other therapies** | **Monitor and follow-up** |
|---|---|---|---|---|
| Pneumonitis | Grade 2 | Withhold | • Administer corticosteroids (initial dose of 1-2 mg/kg prednisone or equivalent) followed by taper | • Monitor participants for signs and symptoms of pneumonitis<br>• Evaluate participants with suspected pneumonitis with radiographic imaging and initiate corticosteroid treatment<br>• Add prophylactic antibiotics for opportunistic infections |
| | Grade 3 or 4, or recurrent Grade 2 | Permanently discontinue | | |
| Diarrhea / Colitis | Grade 2 or 3 | Withhold | • Administer corticosteroids (initial dose of 1-2 mg/kg prednisone or equivalent) followed by taper | • Monitor participants for signs and symptoms of enterocolitis (ie, diarrhea, abdominal pain, blood or mucus in stool with or without fever) and of bowel perforation (ie, peritoneal signs and ileus).<br>• Participants with ≥ Grade 2 diarrhea suspecting colitis should consider GI consultation and performing endoscopy to rule out colitis.<br>• Participants with diarrhea/colitis should be advised to drink liberal quantities of clear fluids. If sufficient oral fluid intake is not feasible, fluid and electrolytes should be substituted via IV infusion. |
| | Grade 4 | Permanently discontinue | | |

| | | | | |
|---|---|---|---|---|
| AST / ALT elevation or Increased bilirubin | Grade 2 | Withhold | • Administer corticosteroids (initial dose of 0.5- 1 mg/kg prednisone or equivalent) followed by taper | • Monitor with liver function tests (consider weekly or more frequently until liver enzyme value returned to baseline or is stable |
| | Grade 3 or 4 | Permanently discontinue | • Administer corticosteroids (initial dose of 1-2 mg/kg prednisone or equivalent) followed by taper | |
| Type 1 diabetes mellitus (T1DM) or Hyperglycemia | Newly onset T1DM or Grade 3 or 4 hyperglycemia associated with evidence of β-cell failure | Withhold | • Initiate insulin replacement therapy for participants with T1DM<br>• Administer anti-hyperglycemic in participants with hyperglycemia | • Monitor participants for hyperglycemia or other signs and symptoms of diabetes. |
| Hypophysitis | Grade 2 | Withhold | • Administer corticosteroids and initiate hormonal replacements as clinically indicated. | • Monitor for signs and symptoms of hypophysitis (including hypopituitarism and adrenal insufficiency) |
| | Grade 3 or 4 | Withhold or permanently discontinue[1] | | |
| Hyperthyroidism | Grade 2 | Continue | • Treat with non-selective beta-blockers (eg, propranolol) or thionamides as appropriate | • Monitor for signs and symptoms of thyroid disorders. |
| | Grade 3 or 4 | Withhold or permanently discontinue[1] | | |
| Hypothyroidism | Grade 2-4 | Continue | • Initiate thyroid replacement hormones (eg, levothyroxine or liothyroinine) per standard of care | • Monitor for signs and symptoms of thyroid disorders. |
| Nephritis and Renal dysfunction | Grade 2 | Withhold | • Administer corticosteroids (prednisone 1-2 mg/kg or equivalent) followed by taper. | • Monitor changes of renal function |
| | Grade 3 or 4 | Permanently discontinue | | |
| Myocarditis | Grade 1 or 2 | Withhold | • Based on severity of AE administer corticosteroids | • Ensure adequate evaluation to confirm etiology and/or exclude other causes |
| | Grade 3 or 4 | Permanently discontinue | | |

| | | | | |
|---|---|---|---|---|
| All other immune-related AEs | Intolerable/ persistent Grade 2 | Withhold | • Based on type and severity of AE administer corticosteroids | • Ensure adequate evaluation to confirm etiology and/or exclude other causes |
| | Grade 3 | Withhold or discontinue based on the type of event. Events that require discontinuation include and not limited to: Gullain-Barre Syndrome, encephalitis | | |
| | Grade 4 or recurrent Grade 3 | Permanently discontinue | | |
| 1. Withhold or permanently discontinue pembrolizumab is at the discretion of the investigator or treating physician.<br>**NOTE:**<br>For participants with Grade 3 or 4 immune-related endocrinopathy where withhold of pembrolizumab is required, pembrolizumab may be resumed when AE resolves to ≤ Grade 2 and is controlled with hormonal replacement therapy or achieved metabolic control (in case of T1DM). | | | | |

In case toxicity does not resolve to Grade 0-1 within 6 weeks after last infusion, trial treatment should be discontinued after consultation with the Sponsor and Primary Investigator. With investigator and Sponsor agreement, subjects with a laboratory adverse event still at Grade 2 after 6 weeks may continue treatment in the trial only if asymptomatic and controlled. For information on the management of adverse events, see Section 5.6.1.

Subjects who experience a recurrence of the same severe or life-threatening event at the same grade or greater with re-challenge of pembrolizumab should be discontinued from trial treatment.

##### 5.2.2 Timing of Dose Administration

Trial treatment should be administered on Days 1 and 8 of each cycle after all procedures/assessments have been completed as detailed on the Trial Flow Chart (Section 6.0). Trial treatment may be administered up to 3 days before or after the scheduled Day 1 of each cycle due to administrative reasons.

All trial treatments will be administered on an outpatient basis.

Gemcitabine should be infused prior to pembrolizumab infusion, according to institutional standard protocol. Antiemetics should be administered according to institutional standard of care protocol, with the exception of the prohibition of prophylactic corticosteroids as antiemetic (See Section 5.6.1 regarding supportive care medications).

Pembrolizumab will be administered as a 30 minute IV infusion (treatment cycle intervals may be increased due to toxicity as described in Section 5.2.1.2). Sites should make every effort to target infusion timing to be as close to 30 minutes as possible. However, given the variability of infusion pumps from site to site, a window of -5 minutes and +10 minutes is permitted (i.e., infusion time is 30 minutes: -5 min/+10 min).

The Procedures Manual contains specific instructions for pembrolizumab dose calculation, reconstitution, preparation of the infusion fluid, and administration.

##### 5.2.3 Trial Blinding/Masking

This is an open-label non-randomized trial; therefore, the Sponsor, investigator and subject will know the treatments administered.

#### 5.3 Randomization or Treatment Allocation

N/A

#### 5.4 Stratification

No stratification will be used for this study.

#### 5.5 Concomitant Medications/Vaccinations (allowed & prohibited)

Medications or vaccinations specifically prohibited in the exclusion criteria are not allowed during the ongoing trial. If there is a clinical indication for one of these or other medications or vaccinations specifically prohibited during the trial, discontinuation from trial therapy or vaccination may be required. The investigator should discuss any questions regarding this with the Sponsor. The final decision on any supportive therapy or vaccination rests with the investigator and/or the subject's primary physician. However, the decision to continue the subject on trial therapy or vaccination schedule requires the mutual agreement of the Primary Investigator, the Sponsor, and the subject.

##### 5.5.1 Acceptable Concomitant Medications

All treatments that the investigator considers necessary for a subject's welfare may be administered at the discretion of the investigator in keeping with the community standards of medical care. All concomitant medication will be recorded on the case report form (CRF) including all prescription, over-the-counter (OTC), herbal supplements, and IV medications and fluids. If changes occur during the trial period, documentation of drug dosage, frequency, route, and date may also be included on the CRF.

All concomitant medications received within 28 days before the first dose of trial treatment and 30 days after the last dose of trial treatment should be recorded. Concomitant medications administered after 30 days after the last dose of trial treatment should be recorded for SAEs and ECIs as defined in Section 7.2.

##### 5.5.2 Prohibited Concomitant Medications

Subjects are prohibited from receiving the following therapies during the Screening and Treatment Phase (including retreatment for post-complete response relapse) of this trial:

- Anti-cancer systemic chemotherapy or biological therapy
- Immunotherapy not specified in this protocol
- Chemotherapy not specified in this protocol
- Investigational agents other than pembrolizumab and gemcitabine
- Radiation therapy
    - Note: Radiation therapy to a symptomatic solitary lesion or to the brain may be allowed after consultation with Sponsor and Primary Investigator.
- Live vaccines within 30 days prior to the first dose of trial treatment and while participating in the trial. Examples of live vaccines include, but are not limited to, the following: measles, mumps, rubella, chicken pox, yellow fever, rabies, BCG, and typhoid (oral) vaccine. Seasonal influenza vaccines for injection are generally killed virus vaccines and are allowed; however intranasal influenza vaccines (e.g. Flu-Mist®) are live attenuated vaccines, and are not allowed.
- Glucocorticoids for any purpose other than to modulate symptoms from an event of clinical interest of suspected immunologic etiology. The use of physiologic doses of corticosteroids may be approved after consultation with the Sponsor and Primary Investigator.

Subjects who, in the assessment by the investigator, require the use of any of the aforementioned treatments for clinical management should be removed from the trial. Subjects may receive other medications that the investigator deems to be medically necessary.

The Exclusion Criteria describes other medications which are prohibited in this trial.

There are no prohibited therapies during the Post-Treatment Follow-up Phase.

#### 5.6 Rescue Medications & Supportive Care

##### 5.6.1 Supportive Care Guidelines

Subjects should receive appropriate supportive care measures (including institutional standard of care supportive measures such as antiemetic prophylaxis prior to chemotherapy administration) as deemed necessary by the treating investigator including but not limited to the items outlined below:

- Diarrhea: Subjects should be carefully monitored for signs and symptoms of enterocolitis (such as diarrhea, abdominal pain, blood or mucus in stool, with or without fever) and of bowel perforation (such as peritoneal signs and ileus). In symptomatic subjects, infectious etiologies should be ruled out, and if symptoms are persistent and/or severe, endoscopic evaluation should be considered.
  - In subjects with severe enterocolitis (Grade 3), pembrolizumab will be permanently discontinued and treatment with systemic corticosteroids should be initiated at a dose of 1 to 2 mg/kg/day of prednisone or equivalent. When symptoms improve to Grade 1 or less, corticosteroid taper should be started and continued over at least 1 month.
  - In subjects with moderate enterocolitis (Grade 2), pembrolizumab should be withheld and anti-diarrheal treatment should be started. If symptoms are persistent for more than one week, systemic corticosteroids should be initiated (e.g., 0.5 mg/kg/day of prednisone or equivalent). When symptoms improve to Grade 1 or less, corticosteroid taper should be started and continued over at least 1 month. Regarding guidelines for continuing treatment with pembrolizumab, see Section 5.2.
  - All subjects who experience diarrhea should be advised to drink liberal quantities of clear fluids. If sufficient oral fluid intake is not feasible, fluid and electrolytes should be substituted via IV infusion.
- Nausea/vomiting: Nausea and vomiting should be treated aggressively, including administration of prophylactic antiemetic therapy according to standard institutional practice. The use of corticosteroids as antiemetic prophylaxis is prohibited. Subjects should be strongly encouraged to maintain liberal oral fluid intake.
- Anti-infectives: Subjects with a documented infectious complication should receive oral or IV antibiotics or other anti-infective agents as considered appropriate by the treating investigator for a given infectious condition, according to standard institutional practice.

- Immune-related adverse events: Please see Section 5.6.1.1 below and the separate guidance document in the administrative binder regarding diagnosis and management of adverse experiences of a potential immunologic etiology.

- Management of Infusion Reactions: Acute infusion reactions (which can include cytokine release syndrome, angioedema, or anaphylaxis) are different from allergic/hypersensitive reactions, although some of the manifestations are common to both AEs. Signs and symptoms usually develop during or shortly after drug infusion and generally resolve completely within 24 hours of completion of infusion. Signs/symptoms may include: Allergic reaction/hypersensitivity (including drug fever); Arthralgia (joint pain); Bronchospasm; Cough; Dizziness; Dyspnea (shortness of breath); Fatigue (asthenia, lethargy, malaise); Headache; Hypertension; Hypotension; Myalgia (muscle pain); Nausea; Pruritis/itching; Rash/desquamation; Rigors/chills; Sweating (diaphoresis); Tachycardia; Tumor pain (onset or exacerbation of tumor pain due to treatment); Urticaria (hives, welts, wheals); Vomiting.

Table 6 below shows treatment guidelines for subjects who experience an infusion reaction associated with administration of pembrolizumab.

Table 6 Infusion Reaction Treatment Guidelines

| **NCI CTCAE Grade** | **Treatment** | **Premedication at subsequent dosing** |
|---|---|---|
| <u>Grade 1</u><br>Mild reaction; infusion interruption not indicated; intervention not indicated | Increase monitoring of vital signs as medically indicated until the subject is deemed medically stable in the opinion of the investigator. | None |
| <u>Grade 2</u><br>Requires infusion interruption but responds promptly to symptomatic treatment (e.g., antihistamines, NSAIDS, narcotics, IV fluids); prophylactic medications indicated for < =24 hrs | **Stop Infusion and monitor symptoms.**<br>Additional appropriate medical therapy may include but is not limited to:<br>IV fluids<br>Antihistamines<br>NSAIDS<br>Acetaminophen<br>Narcotics<br>Increase monitoring of vital signs as medically indicated until the subject is deemed medically stable in the opinion of the investigator.<br>If symptoms resolve within one hour of stopping drug infusion, the infusion may be restarted at 50% of the original infusion rate (e.g. from 100 mL/hr to 50 mL/hr). Otherwise dosing will be held until symptoms resolve and the subject should be premedicated for the next scheduled dose.<br>**Subjects who develop Grade 2 toxicity despite adequate premedication should be permanently discontinued from further trial treatment administration.** | Subject may be premedicated 1.5h (± 30 minutes) prior to infusion of pembrolizumab with:<br><br>Diphenhydramine 50 mg po (or equivalent dose of antihistamine).<br><br>Acetaminophen 500-1000 mg po (or equivalent dose of antipyretic). |

| NCI CTCAE Grade | Treatment | Premedication at subsequent dosing |
|---|---|---|
| Grades 3 or 4<br>Grade 3:<br>Prolonged (i.e., not rapidly responsive to symptomatic medication and/or brief interruption of infusion); recurrence of symptoms following initial improvement; hospitalization indicated for other clinical sequelae (e.g., renal impairment, pulmonary infiltrates)<br><br>Grade 4:<br>Life-threatening; pressor or ventilatory support indicated | **Stop Infusion.**<br>Additional appropriate medical therapy may include but is not limited to:<br>IV fluids<br>Antihistamines<br>NSAIDS<br>Acetaminophen<br>Narcotics<br>Oxygen<br>Pressors<br>Corticosteroids<br>Epinephrine<br><br>Increase monitoring of vital signs as medically indicated until the subject is deemed medically stable in the opinion of the investigator.<br>Hospitalization may be indicated.<br>**Subject is permanently discontinued from further trial treatment administration.** | No subsequent dosing |
| Appropriate resuscitation equipment should be available in the room and a physician readily available during the period of drug administration.<br>For Further information, please refer to the Common Terminology Criteria for Adverse Events v4.0 (CTCAE) at http://ctep.cancer.gov | | |

###### 5.6.1.1 Supportive Care Guidelines for Events of Clinical Interest and Immune-related Adverse Events (irAEs)

Events of clinical interest of a potential immunologic etiology (irECIs) may be defined as an adverse event of unknown etiology, associated with drug exposure and is consistent with an immune phenomenon. irAEs may be predicted based on the nature of the pembrolizumab compound, its mechanism of action, and reported experience with immunotherapies that have a similar mechanism of action. Special attention should be paid to AEs that may be suggestive of potential irAEs. An irAE can occur shortly after the first dose or several months after the last dose of treatment.

If an irAE is suspected, efforts should be made to rule out neoplastic, infectious, metabolic, toxin or other etiologic causes prior to labeling an adverse event as an irAE. Information on how to identify and evaluate irAEs has been developed and is included in the Event of Clinical Interest and Immune-Related Adverse Event Guidance Document located in the Administrative Binder. Subjects who develop a Grade 2 or higher irAE should be discussed immediately with the Sponsor and Primary Investigator.

Recommendations to managing irAEs not detailed elsewhere in the protocol are detailed in Table 7.

Table 7 General Approach to Handling irAEs

| irAE | Withhold/Discontinue pembrolizumab? | Supportive Care |
|---|---|---|
| Grade 1 | No action | Provide symptomatic treatment |
| Grade 2 | May withhold pembrolizumab | Consider systemic corticosteroids in addition to appropriate symptomatic treatment |
| Grade 3 and Grade 4 | Withhold pembrolizumab<br>Discontinue if unable to reduce corticosteroid dose to < 10 mg per day prednisone equivalent within 12 weeks of toxicity | Systemic corticosteroids are indicated in addition to appropriate symptomatic treatment. May utilize 1 to 2 mg/kg prednisone or equivalent per day.<br>Steroid taper should be considered once symptoms improve to Grade 1 or less and tapered over at least 4 weeks. |

##### 5.6.1.2 Supportive Care Guidelines for Pneumonitis

Subjects with symptomatic pneumonitis should immediately stop receiving pembrolizumab and have an evaluation. The evaluation may include chest radiography, bronchoscopy, and pulmonary function tests to rule out other causes such as infection. If the subject is determined to have study drug associated pneumonitis, the suggested treatment plan is detailed in Table 8.

Table 8. Recommended Approach to Handling Pneumonitis

| Study drug associated pneumonitis | Withhold/Discontinue pembrolizumab? | Supportive Care |
|---|---|---|
| Grade 1 (asymptomatic) | No action | Intervention not indicated |
| Grade 2 | Withhold pembrolizumab, may return to treatment if improves to Grade 1 or resolves within 12 weeks<br>Report immediately as ECI | Consider pulmonary consultation with bronchoscopy and biopsy/BAL and Infectious Disease consultation as appropriate.<br>Systemic corticosteroids are indicated (1-2 mg/kg daily prednisone or equivalent). Patients with suspected or diagnosed pneumonitis should be started on steroid treatment immediately, and steroid treatment should not be delayed for a therapeutic trial of antibiotics. Treatment of both a potential infectious etiology and pneumonitis in parallel may be warranted.<br>When symptoms improve to Grade 1 or less, steroid taper should be started and continued over no less than 4 weeks. Permanently discontinue for inability to reduce corticosteroid dose to 10 mg or less of prednisone or equivalent per day within 12 weeks.<br>Conduct an in person evaluation approximately twice weekly until the patient is improving.<br>Consider frequent chest x-ray as part of monitoring.<br>Discontinue pembrolizumab if upon re-challenge the patient develops a second episode of Grade 2 or higher pneumonitis. |

| | | |
|---|---|---|
| Grade 3 and Grade 4 | Permanently discontinue pembrolizumab<br><br>Report immediately as ECI. | Hospitalize patient and consider bronchoscopy with biopsy and/or BAL.<br><br>Systemic corticosteroids are indicated (methylprednisolone 125 mg IV). When symptoms improve to Grade 1 or less, a high dose oral steroid (prednisone 1-2 mg/kg daily or dexamethasone 4 mg every 4 hours) taper should be started and continued over no less than 4 weeks. If IV steroids followed by high dose oral steroids does not reduce initial symptoms within 48 to 72 hours, treat with additional anti-inflammatory measures.<br><br>Discontinue anti-inflammatory measures upon symptom relief and initiate a prolonged steroid taper over 45 to 60 days. If symptoms worsen during steroid reduction, initiate a re-tapering of steroids starting at a higher dose of 80 or 100 mg followed by a more prolonged taper and administer additional anti-inflammatory measures, as needed. In addition, prophylactic antibiotics for opportunistic infections should be considered.<br><br>The use of infliximab may be indicated as appropriate.<br><br>Refer to the Event of Clinical Interest and Immune-related Adverse Event Guidance Document for additional recommendations. |

For Grade 2 pneumonitis that improves to ≤ Grade 1 within 12 weeks, the following rules should apply:

- First episode of pneumonitis
  - May increase dosing interval by one week in subsequent cycles
- Second episode of pneumonitis – permanently discontinue pembrolizumab if upon rechallenge subject develops pneumonitis ≥ Grade 2

#### 5.7 Diet/Activity/Other Considerations

##### 5.7.1 Diet

Subjects should maintain a normal diet unless modifications are required to manage an AE such as diarrhea, nausea or vomiting.

##### 5.7.2 Contraception

Pembrolizumab and gemcitabine may have adverse effects on a fetus in utero. Furthermore, it is not known if pembrolizumab has transient adverse effects on the composition of sperm. Non-pregnant, non-breast-feeding women may be enrolled if they are willing to use 2 methods of birth control or are considered highly unlikely to conceive. Highly unlikely to conceive is defined as 1) surgically sterilized, or 2) postmenopausal (a woman who is ≥45 years of age and has not had menses for greater than 1 year will be considered postmenopausal), or 3) not heterosexually active for the duration of the study. The two birth control methods can be either two barrier methods or a barrier method plus a hormonal method to prevent pregnancy. Subjects should start using birth control from study Visit 1 throughout the study period up to 120 days after the last dose of study therapy.

The following are considered adequate barrier methods of contraception: diaphragm, condom (by the partner), copper intrauterine device, sponge, or spermicide. Appropriate hormonal contraceptives will include any registered and marketed contraceptive agent that contains an estrogen and/or a progestational agent (including oral, subcutaneous, intrauterine, or intramuscular agents).

Subjects should be informed that taking the study medications may involve unknown risks to the fetus (unborn baby) if pregnancy were to occur during the study. In order to participate in the study they must adhere to the contraception requirement (described above) for the duration of the study and during the follow-up period defined in section 7.2.2-Reporting of Pregnancy and Lactation to the Primary Investigator, and to Merck. If there is any question that a subject will not reliably comply with the requirements for contraception, that subject should not be entered into the study.

##### 5.7.3 Use in Pregnancy

If a subject inadvertently becomes pregnant while on treatment, the subject will immediately be removed from the study. The site will contact the subject at least monthly and document the subject's status until the pregnancy has been completed or terminated. The outcome of the pregnancy will be reported to the Primary Investigator and to Merck without delay and within 24 hours if the outcome is a serious adverse experience (e.g., death, abortion, congenital anomaly, or other disabling or life-threatening complication to the mother or newborn). The study investigator will make every effort to obtain permission to follow the outcome of the pregnancy and report the condition of the fetus or newborn to the Sponsor. If a male subject impregnates his female partner the study personnel at the site must be informed immediately and the pregnancy reported to the Primary Investigator and to Merck and followed as described above and in Section 7.2.2.

##### 5.7.4 Use in Nursing Women

It is unknown whether pembrolizumab is excreted in human milk. Significant risk to the infant via human milk exposure to gemcitabine or pembrolizumab cannot be ruled out. Since many drugs are excreted in human milk, and because of the potential for serious adverse reactions in the nursing infant, subjects who are breast-feeding are not eligible for enrollment.

#### 5.8 Subject Withdrawal/Discontinuation Criteria

Subjects may withdraw consent at any time for any reason or be discontinued from the trial at the discretion of the investigator should any untoward effect occur. In addition, a subject may be withdrawn by the investigator or the Sponsor if enrollment into the trial is inappropriate, the trial plan is violated, or for administrative and/or other safety reasons. Specific details regarding discontinuation or withdrawal are provided in Section 7.1.4 – Other Procedures.

A subject must be discontinued from the trial for any of the following reasons:

- The subject or legal representative (such as a parent or legal guardian) withdraws consent.

- Confirmed radiographic disease progression

  *Note*: Subjects who are clinically stable or who are clinically benefitting from therapy may continue treatment with pembrolizumab past first progression on imaging, until confirmation of disease progression on second imaging. Gemcitabine will not be continued beyond first radiographic progression (RECIST v1.1, see Section 12.3).

  *Note*: A subject may be granted an exception to continue on treatment with confirmed radiographic progression if clinically stable or clinically improved.

- Unacceptable adverse experiences as described in Section 5.2.1.2

- Intercurrent illness that prevents further administration of treatment

- Investigator's decision to withdraw the subject

- The subject has a confirmed positive serum pregnancy test

- Noncompliance with trial treatment or procedure requirements

- The subject is lost to follow-up

- Completed 24 months of treatment with pembrolizumab

  *Note: 24 months of study medication is calculated from the date of first dose. Subjects who stop pembrolizumab after 24 months may be eligible for up to one year of additional study treatment if they progress after stopping study treatment based upon approval of the Primary Investigator and Merck, including review of clinical and laboratory status.*

- Administrative reasons

The End of Treatment and Follow-up visit procedures are listed in Section 6 (Protocol Flow Chart) and Section 7.1.5 (Visit Requirements). After the end of treatment, each subject will be followed for 30 days for adverse event monitoring (serious adverse events will be collected for 90 days after the end of treatment as described in Section 7.2.3.1). Subjects who discontinue for reasons other than progressive disease will have post-treatment follow-up for disease status until disease progression, initiating a non-study cancer treatment, withdrawing consent or becoming lost to follow-up. After documented disease progression each subject will be followed by telephone for overall survival until death, withdrawal of consent, or the end of the study, whichever occurs first.

#### 5.9 Subject Replacement Strategy

Subjects in the initial phase I safety portion of the study who are not able to complete the evaluation period for safety will be replaced.

#### 5.10 Clinical Criteria for Early Trial Termination

Early trial termination will be the result of the criteria specified below:

1. Quality or quantity of data recording is inaccurate or incomplete

2. Poor adherence to protocol and regulatory requirements

3. Incidence or severity of adverse drug reaction in this or other studies indicates a potential health hazard to subjects

4. Plans to modify or discontinue the development of the study drug

In the event of Merck decision to no longer supply study drug, ample notification will be provided so that appropriate adjustments to subject treatment can be made.

#### 6.0 TRIAL FLOW CHART

#### 6.1 Study Flow Chart

Baseline evaluations are to be conducted within 4 weeks prior to enrollment except for EKG which may be performed within 8 weeks prior to enrollment. Scans must be done within 4 weeks prior to the start of therapy.

| **Trial Period:** | **Screening Phase** | | **Treatment Cycles** | | | | | | | | | | | | **End of Treatment** | **Post-Treatment** | | |
|---|---|---|---|---|---|---|---|---|---|---|---|---|---|---|---|---|---|---|
| Treatment Cycle/Title: | Pre-screening (Visit 1) | Main Study Screening (Visit 2) | C1/ D1 | C1/ D8 | C2 /D1 | C2/ D8 | C3/ D1 | C3/ D8 | C4/ D1 | C4/ D8 | To be repeated beyond 8 cycles 5 | 6 | 7 | 8 | Discon | Safety Follow -up | Follo w Up Visits | Surviva l Follow-Up |
| Scheduling Window (Days): | | -28 to -1 | | | ± 3 | | ± 3 | | ± 3 | | ± 3 | ± 3 | ± 3 | ± 3 | At time of Discon | 30 days post discon | Every 8 weeks post discon | Every 12 weeks |
| | | **Administrative Procedures** | | | | | | | | | | | | | | | | |
| Informed Consent | X | | | | | | | | | | | | | | | | | |
| Inclusion/Exclusion Criteria | X | | | | | | | | | | | | | | | | | |
| Demographics and Medical History | X | | | | | | | | | | | | | | | | | |
| Prior and Concomitant Medication Review | X | X | X | | X | | X | | X | | X | X | X | X | X | X | X | X |
| Pembrolizumab Administration[a] | | | X | | X | | X | | X | | X | X | X | X | | | | |
| Gemcitabine Administration[b] | | | X | X | X | X | X | X | X | X | X (Days 1, 8 for a maximum of 6 cycles) | | | | | | | |

| **Trial Period:** | **Screening Phase** | | **Treatment Cycles** | | | | | | | | | | | | **End of Treatment** | **Post-Treatment** | | |
|---|---|---|---|---|---|---|---|---|---|---|---|---|---|---|---|---|---|---|
| | | | | | | | | | | | To be repeated beyond 8 cycles | | | | | | | |
| Treatment Cycle/Title: | Pre-screening (Visit 1) | Main Study Screening (Visit 2) | C1/ D1 | C1/ D8 | C2 /D1 | C2/ D8 | C3/ D1 | C3/ D8 | C4/ D1 | C4/ D8 | 5 | 6 | 7 | 8 | Discon | Safety Follow -up | Follo w Up Visits | Surviva l Follow-Up |
| Scheduling Window (Days): | | -28 to -1 | | | ± 3 | | ± 3 | | ± 3 | | ± 3 | ± 3 | ± 3 | ± 3 | At time of Discon | 30 days post discon | Every 8 weeks post discon | Every 12 weeks |
| Post-study anticancer therapy status | | | | | | | | | | | | | | | | | X | X |
| Survival Status | | | | | | | | | | | | | | | X | X | X | X |
| | | **Clinical Procedures/Assessments** | | | | | | | | | | | | | | | | |
| Review Adverse Events | | | X | X | X | X | X | X | X | X | X | X | X | X | X | X | X | |
| Full Physical Examination | | X | X | | X | | X | | X | | X | X | X | X | X | X | | |
| Vital Signs and Weight | X | X | X | X | X | X | X | X | X | X | X | X | X | X | X | X | X | |
| ECOG Performance Status | X | X | X | | X | | X | | X | | X | X | X | X | X | X | X | |
| EKG | | X | | | | | | | | | | | | | | | | |
| | | **Laboratory Procedures/Assessments: analysis performed by LOCAL laboratory** | | | | | | | | | | | | | | | | |
| HIV, Hepatitis serologies | | X[c] | | | | | | | | | | | | | | | | |
| Pregnancy Test – Urine or Serum β-HCG | | X[d] | | | X[d] | | X[d] | | X[d] | | X[d] | X[d] | X[d] | X[d] | | | | |
| PT/INR and aPTT | | X | | | | | | | | | | | | | | | | |
| CBC with Differential | | X | X | X | X | X | X | X | X | X | X (Days 1 and 8 through | X | X | X | X | X | | |

| **Trial Period:** | **Screening Phase** | | **Treatment Cycles** | | | | | | | | | | | | **End of Treatment** | **Post-Treatment** | | |
|---|---|---|---|---|---|---|---|---|---|---|---|---|---|---|---|---|---|---|
| | | | | | | | | | | | To be repeated beyond 8 cycles | | | | | | | |
| Treatment Cycle/Title: | Pre-screening (Visit 1) | Main Study Screening (Visit 2) | C1/ D1 | C1/ D8 | C2 /D1 | C2/ D8 | C3/ D1 | C3/ D8 | C4/ D1 | C4/ D8 | 5 | 6 | 7 | 8 | Discon | Safety Follow -up | Follo w Up Visits | Surviva l Follow-Up |
| Scheduling Window (Days): | | -28 to -1 | | | ± 3 | | ± 3 | | ± 3 | | ± 3 | ± 3 | ± 3 | ± 3 | At time of Discon | 30 days post discon | Every 8 weeks post discon | Every 12 weeks |
| | | | | | | | | | | | cycle 6 gemcitabine ) | | | | | | | |
| Comprehensive Serum Chemistry Panel[e] | | X | X | | X | | X | | X | | X | X | X | X | X | X | | |
| Urinalysis | | X | X | | X | | X | | X | | X | X | X | X | | | | |
| T3, FT4 and TSH[f] | | X | X | | | | | | X | | | X | | | | | | |
| | | **Efficacy Measurements** | | | | | | | | | | | | | | | | |
| Tumor Imaging (CT Thorax including entire liver and adrenal glands) | X | | | | | | X[g] | | | | X[g] | | | | | | | X |
| | | **Archival Tissue Collection/Correlative Studies Blood** | | | | | | | | | | | | | | | | |
| Blood for immune parameters | | | X | | | | | | | | | X | | | | | | X[h] |
| ProtoArray® Immune Response Biomarker Profiling | | | X | | | | | | | | | X | | | | | | |
| Submit archived slides (if available) | | X | | | | | | | | | | | | | | | | |

| **Trial Period:** | **Screening Phase** | | **Treatment Cycles** | | | | | | | | | | | | **End of Treatment** | **Post-Treatment** | | |
|---|---|---|---|---|---|---|---|---|---|---|---|---|---|---|---|---|---|---|
| | | | | | | | | | | | To be repeated beyond 8 cycles | | | | | | | |
| Treatment Cycle/Title: | Pre-screening (Visit 1) | Main Study Screening (Visit 2) | C1/ D1 | C1/ D8 | C2 /D1 | C2/ D8 | C3/ D1 | C3/ D8 | C4/ D1 | C4/ D8 | 5 | 6 | 7 | 8 | Discon | Safety Follow -up | Follo w Up Visits | Surviva l Follow-Up |
| Scheduling Window (Days): | | -28 to -1 | | | ± 3 | | ± 3 | | ± 3 | | ± 3 | ± 3 | ± 3 | ± 3 | At time of Discon | 30 days post discon | Every 8 weeks post discon | Every 12 weeks |
| for immunoscore and PD-L1 testing | | | | | | | | | | | | | | | | | | |

a. Pembrolizumab administration will be every 21 days for 2 years, or until disease progression or prohibitive toxicity, whichever comes first.
b. Gemcitabine will be administered on Days 1 and 8 of each 21 day cycle for a maximum of 6 cycles, or until disease progression or prohibitive toxicity, whichever comes first.
c. Within 6 months of enrollment
d. All females with childbearing potential starting at baseline, and then with Day 1 of each cycle throughout pembrolizumab administration.
e. Albumin, Alkaline Phosphatase, Total bilirubin, Bicarbonate, BUN, Calcium, Chloride, Creatinine, Glucose, LDH, Phosphorus, Potassium, Total Protein, SGOT [AST], SGPT [ALT], Sodium
f. TSH, free T3, free T4 should continue every 6 weeks beginning at Cycle 6 of study treatment and continue throughout study treatment.
g. CT scan must be done prior to Week 7 treatment. Disease assessment (CT scans) should be performed every 6 weeks for 2 years, then every 2 months while in follow up. Imaging to include pelvis or other locations as clinically appropriate.
h. Every 12 weeks for 2 years or until progressive disease.

### 7.0 TRIAL PROCEDURES

#### 7.1 Trial Procedures

The Trial Flow Chart - Section 6.0 summarizes the trial procedures to be performed at each visit. Individual trial procedures are described in detail below. It may be necessary to perform these procedures at unscheduled time points if deemed clinically necessary by the investigator.

Furthermore, additional evaluations/testing may be deemed necessary by the Investigator and/or Merck for reasons related to subject safety. In some cases, such evaluation/testing may be potentially sensitive in nature (e.g., HIV, Hepatitis C, etc.), and thus local regulations may require that additional informed consent be obtained from the subject. In these cases, such evaluations/testing will be performed in accordance with those regulations.

##### 7.1.1 Administrative Procedures

###### 7.1.1.1 Informed Consent

The Investigator must obtain documented consent from each potential subject prior to participating in a clinical trial.

###### 7.1.1.1.1 General Informed Consent

Consent must be documented by the subject's dated signature or by the subject's legally acceptable representative's dated signature on a consent form along with the dated signature of the person conducting the consent discussion.

A copy of the signed and dated consent form should be given to the subject before participation in the trial.

The initial informed consent form, any subsequent revised written informed consent form and any written information provided to the subject must receive the IRB/ERC's approval/favorable opinion in advance of use. The subject or his/her legally acceptable representative should be informed in a timely manner if new information becomes available that may be relevant to the subject's willingness to continue participation in the trial. The communication of this information will be provided and documented via a revised consent form or addendum to the original consent form that captures the subject's dated signature or by the subject's legally acceptable representative's dated signature.

Specifics about a trial and the trial population will be added to the consent form template at the protocol level.

The informed consent will adhere to IRB/ERC requirements, applicable laws and regulations and Sponsor requirements.

###### 7.1.1.2 Inclusion/Exclusion Criteria

All inclusion and exclusion criteria will be reviewed by the investigator or qualified designee to ensure that the subject qualifies for the trial.

###### 7.1.1.3 Subject Identification Card

Not applicable.

###### 7.1.1.4 Medical History

A medical history will be obtained by the investigator or qualified designee. Medical history will include all active conditions, and any condition diagnosed within the prior 10 years that are considered to be clinically significant by the Investigator. Details regarding the disease for which the subject has enrolled in this study will be recorded separately and not listed as medical history.

###### 7.1.1.5 Prior and Concomitant Medications Review

###### 7.1.1.5.1 Prior Medications

The investigator or qualified designee will review prior medication use, including any protocol-specified washout requirement, and record prior medication taken by the subject within 28 days before starting the trial. Treatment for the disease for which the subject has enrolled in this study will be recorded separately and not listed as a prior medication.

###### 7.1.1.5.2 Concomitant Medications

The investigator or qualified designee will record medication, if any, taken by the subject during the trial. All medications related to reportable SAEs and ECIs should be recorded as defined in Section 7.2.

###### 7.1.1.6 Disease Details and Treatments

###### 7.1.1.6.1 Disease Details

The investigator or qualified designee will obtain prior and current details regarding disease status.

###### 7.1.1.6.2 Prior Treatment Details

The investigator or qualified designee will review all prior cancer treatments including systemic treatments, radiation and surgeries.

###### 7.1.1.6.3 Subsequent Anti-Cancer Therapy Status

The investigator or qualified designee will review all new anti-neoplastic therapy initiated after the last dose of trial treatment. If a subject initiates a new anti-cancer therapy within 30 days after the last dose of trial treatment, the 30 day Safety Follow-up visit must occur before the first dose of the new therapy. Once new anti-cancer therapy has been initiated the subject will move into survival follow-up.

###### 7.1.1.7 Assignment of Study Number

Study Numbers will be assigned at enrollment based on order of enrollment and study ID, as follows: MK-01, MK-02, MK-03, etc.

All case report forms, study reports, and laboratory samples for research tests will be labeled with the full patient Study Number.

###### 7.1.1.8 Assignment of Randomization Number

Not applicable

###### 7.1.1.9 Trial Compliance (Medication/Diet/Activity/Other)

Not applicable

##### 7.1.2 Clinical Procedures/Assessments

###### 7.1.2.1 Adverse Event (AE) Monitoring

The investigator or qualified designee will assess each subject to evaluate for potential new or worsening AEs as specified in the Trial Flow Chart and more frequently if clinically indicated. Adverse experiences will be graded and recorded throughout the study and during the follow-up period according to NCI CTCAE Version 4.0 (see Section 12.2). Toxicities will be characterized in terms regarding seriousness, causality, toxicity grading, and action taken with regard to trial treatment.

All AEs of unknown etiology associated with pembrolizumab exposure should be evaluated to determine if it is possibly an event of clinical interest (ECI) of a potentially immunologic etiology (irAE). See Section 5.6.1.1 and the separate guidance document in the administrative binder regarding the identification, evaluation and management of AEs of a potential immunological etiology.

Please refer to section 7.2 for detailed information regarding the assessment and recording of AEs.

###### 7.1.2.2 Full Physical Exam

The investigator or qualified designee will perform a complete physical exam during the screening period. Clinically significant abnormal findings should be recorded as medical history. A full physical exam should be performed during screening,

###### 7.1.2.3 Directed Physical Exam

Additional directed physical examinations will be performed as deemed appropriate by the investigator or qualified designee in assessment of symptoms or side effects.

###### 7.1.2.4 Vital Signs

The investigator or qualified designee will take vital signs at screening, prior to the administration of each dose of trial treatment and at treatment discontinuation as specified in the Trial Flow Chart (Section 6.0). Vital signs should include temperature, pulse, respiratory rate, pulse oximetry recording, weight and blood pressure. Height will be measured at screening only.

###### 7.1.2.5 Eastern Cooperative Oncology Group (ECOG) Performance Scale

The investigator or qualified designee will assess ECOG status (see Section 12.1) at screening, prior to the administration of each dose of trial treatment and discontinuation of trial treatment as specified in the Trial Flow Chart.

###### 7.1.2.6 Tumor Imaging and Assessment of Disease

Tumor response will be measured using RECIST v1.1 (See Section 12.3). CT scans will be performed every 6 weeks for 2 years, then every 2 months while in follow up.

Subjects with first disease progression may continue to receive pembrolizumab until confirmation of disease progression on next imaging, provided they remain clinically stable and appear to be receiving benefit from therapy. Gemcitabine will not be continued beyond first radiographic progression.

###### 7.1.2.7 Tumor Tissue Collection and Correlative Studies Blood Sampling

Tumor specimens will be evaluated for expression of PD-L1 by immunohistochemistry and for tumor immune score. Immune monitoring will be conducted with collection of peripheral blood and assessment of protein arrays, and compared pre-treatment and at 12 weeks after initiation of pembrolizumab. Additional samples may be archived at 3 month intervals thereafter while receiving pembrolizumab therapy.

Blood for immune parameters: This includes sera for protoArray, and heparinized blood for immunophenotyping and WBC banking. Five x 10cc sodium heparin (green top) tubes and 1 x 10cc serum separator tube (SST) are drawn at the pre-treatment time point and at week 12. Tubes are transported at room temperature to the Immunological Monitoring Laboratory for processing.

##### 7.1.3 Laboratory Procedures/Assessments

Details regarding specific laboratory procedures/assessments to be performed in this trial are provided below. The total amount of blood/tissue to be drawn/collected over the course of the trial (from pre-trial to post-trial visits), including approximate blood/tissue volumes drawn/collected by visit and by sample type per subject can be found in the Procedures Manual.

###### 7.1.3.1 Laboratory Safety Evaluations (Hematology, Chemistry and Urinalysis)

Laboratory tests for hematology, chemistry, urinalysis, and others are specified in Table 9. The total amount of blood/tissue to be drawn/collected over the course of the trial (from pre-trial to post-trial visits), including approximate blood/tissue volumes drawn/collected by visit and by sample type per subject can be found in the Procedures Manual.

Table 9 Laboratory Tests

| Hematology | Chemistry | Urinalysis | Other |
|---|---|---|---|
| Hematocrit | Albumin | Blood | Serum β-human chorionic gonadotropin† |
| Hemoglobin | Alkaline phosphatase | Glucose | (β-hCG)† |
| Platelet count | Alanine aminotransferase (ALT) | Protein | PT (INR) |
| WBC (total and differential) | Aspartate aminotransferase (AST) | Specific gravity | aPTT |
| Red Blood Cell Count | Lactate dehydrogenase (LDH) | Microscopic exam *(If abnormal)* | Total thriiodothyronine (T3) |
| Absolute Neutrophil Count | Carbon Dioxide | results are noted | Free tyroxine (T4) |
| | *($CO_2$ or biocarbonate)* | Urine pregnancy test † | Thyroid stimulating hormone (TSH) |
| | Uric Acid | | |
| | Calcium | | |
| | Chloride | | Blood for correlative studies |
| | Glucose | | |
| | Phosphorus | | |
| | Potassium | | |
| | Sodium | | |
| | Magnesium | | |
| | Total Bilirubin | | |
| | Direct Bilirubin *(If total bilirubin is elevated above the upper limit of normal)* | | |
| | Total protein | | |
| | Blood Urea Nitrogen | | |

† Perform on women of childbearing potential only. If urine pregnancy results cannot be confirmed as negative, a serum pregnancy test will be required.

Laboratory tests for screening should be performed within 10 days prior to the first dose of treatment. After Cycle 1, pre-dose laboratory procedures can be conducted up to 72 hours prior to dosing. Results must be reviewed by the investigator or qualified designee and found to be acceptable prior to each dose of trial treatment.

###### 7.1.3.2 Pharmacokinetic/Pharmacodynamic Evaluations

Not applicable.

##### 7.1.4 Other Procedures

###### 7.1.4.1 Withdrawal/Discontinuation

When a subject discontinues/withdraws prior to trial completion, all applicable activities scheduled for the final trial visit should be performed at the time of discontinuation. Any adverse events which are present at the time of discontinuation/withdrawal should be followed in accordance with the safety requirements outlined in Section 7.2 - Assessing and Recording Adverse Events. Subjects who a) attain a CR or b) complete 24 months of treatment with pembrolizumab may discontinue treatment with the option of restarting treatment if they meet the criteria specified in Section 5.6. After discontinuing treatment following assessment of CR, these subjects should return to the site for a Safety Follow-up Visit (described in Section 7.1.5.3.1) and then proceed to the Follow-Up Period of the study (described in Section 7.1.5.4).

###### 7.1.4.2 Blinding/Unblinding

Not applicable for this study.

##### 7.1.5 Visit Requirements

Visit requirements are outlined in Section 6.0 - Trial Flow Chart. Specific procedure-related details are provided above in Section 7.1 - Trial Procedures.

###### 7.1.5.1 Screening

Baseline evaluations are to be conducted within 4 weeks prior to enrollment except for EKG, which may be performed within 8 weeks prior to enrollment. Scans must be done within 4 weeks prior to the start of therapy.

###### 7.1.5.1.1 Screening Period

During the screening period, subjects will complete procedures as outlined in the Study Flow Chart in Section 6.1. Screening procedures include documentation of informed consent, review and confirmation of eligibility via inclusion and exclusion criteria, review of demographics and medical history, prior and concomitant medication review and documentation, recording of vital signs and weight, ECOG performance status.

Laboratory investigations will include documentation of HIV and hepatitis serologies, pregnancy test (for women of childbearing potential), PT/INR and aPTT, CBC with differential, comprehensive serum chemistry panel, urinalysis, and thyroid studies including T3, free T4, and TSH.

###### 7.1.5.2 Treatment Period

Studies, procedures, and treatments will be performed as outlined in Section 6.1 Study Flow Chart.

###### 7.1.5.3 Post-Treatment Visits

Post-treatment visits will include End of Treatment visit at the time of study discontinuation, Safety follow up visit (as outlined in Section 7.1.5.3.1 below), follow up visits every 8 weeks after study discontinuation.

###### 7.1.5.3.1 Safety Follow-Up Visit

The mandatory Safety Follow-Up Visit should be conducted approximately 30 days after the last dose of trial treatment or before the initiation of a new anti-cancer treatment, whichever comes first. All AEs that occur prior to the Safety Follow-Up Visit should be recorded. Subjects with an AE of Grade > 1 will be followed until the resolution of the AE to Grade 0-1 or until the beginning of a new anti-neoplastic therapy, whichever occurs first. SAEs that occur within 90 days of the end of treatment or before initiation of a new anti-cancer treatment should also be followed and recorded.

###### 7.1.5.4 Follow-up Visits

Subjects who discontinue trial treatment for a reason other than disease progression will move into the Follow-Up Phase and should be assessed every 8 weeks (56 ± 7 days) by radiologic imaging to monitor disease status. Every effort should be made to collect information regarding disease status until the start of new anti-neoplastic therapy, disease progression, death, end of the study or if the subject begins retreatment with pembrolizumab as detailed in Section 5.6. Information regarding post-study anti-neoplastic treatment will be collected if new treatment is initiated.

Subjects who are eligible to receive retreatment with pembrolizumab according to the criteria in Section 5.6 will continued to be followed per protocol until disease progression. Details are provided in Section 6.1 – Trial Flow Chart.

###### 7.1.5.4.1 Survival Follow-up

Once a subject experiences confirmed disease progression or starts a new anti-cancer therapy, the subject moves into the survival follow-up phase and should be contacted by telephone every 12 weeks to assess for survival status until death, withdrawal of consent, or the end of the study, whichever occurs first.

#### 7.2 Assessing and Recording Adverse Events

An adverse event is defined as any untoward medical occurrence in a patient or clinical investigation subject administered a pharmaceutical product and which does not necessarily have to have a causal relationship with this treatment. An adverse event can therefore be any unfavorable and unintended sign (including an abnormal laboratory finding, for example), symptom, or disease temporally associated with the use of a medicinal product or protocol-specified procedure, whether or not considered related to the medicinal product or protocol-specified procedure. Any worsening (i.e., any clinically significant adverse change in frequency and/or intensity) of a preexisting condition that is temporally associated with the use of the pembrolizumab or gemcitabine, is also an adverse event.

Changes resulting from normal growth and development that do not vary significantly in frequency or severity from expected levels are not to be considered adverse events. Examples of this may include, but are not limited to, teething, typical crying in infants and children and onset of menses or menopause occurring at a physiologically appropriate time.

Merck product includes any pharmaceutical product, biological product, device, diagnostic agent or protocol-specified procedure, whether investigational (including placebo or active comparator medication) or marketed, manufactured by, licensed by, provided by or distributed by Merck for human use.

Adverse events may occur during the course of the use of Merck product in clinical trials or within the follow-up period specified by the protocol, or prescribed in clinical practice, from overdose (whether accidental or intentional), from abuse and from withdrawal.

Adverse events may also occur in screened subjects during any pre-allocation baseline period as a result of a protocol-specified intervention, including washout or discontinuation of usual therapy, diet, placebo treatment or a procedure.

All adverse events will be recorded from the time the consent form is signed through 30 days following cessation of treatment and at each examination on the Adverse Event case report forms/worksheets. The reporting timeframe for adverse events meeting any serious criteria is described in section 7.2.3.1.

Adverse events will not be collected for subjects during the pre-screening period (for determination of archival tissue status) as long as that subject has not undergone any protocol-specified procedure or intervention. If the subject requires a blood draw, or any other study procedure, the subject is first required to provide consent to the main study and AEs will be captured according to guidelines for standard AE reporting.

##### 7.2.1 Definition of an Overdose for This Protocol and Reporting of Overdose to the Sponsor and to Merck

For purposes of this trial, an overdose will be defined as any dose exceeding the prescribed dose for gemcitabine or pembrolizumab by 20% over the prescribed dose. No specific information is available on the treatment of overdose of pembrolizumab. In the event of overdose, gemcitabine and pembrolizumab should be discontinued and the subject should be observed closely for signs of toxicity. Appropriate supportive treatment should be provided if clinically indicated.

If an adverse event(s) is associated with ("results from") the overdose of either gemcitabine or a Merck product, the adverse event(s) is reported as a serious adverse event, even if no other seriousness criteria are met.

If a dose of gemcitabine or Merck's product meeting the protocol definition of overdose is taken without any associated clinical symptoms or abnormal laboratory results, the overdose is reported as a non-serious Event of Clinical Interest (ECI), using the terminology "accidental or intentional overdose without adverse effect."

All reports of overdose with and without an adverse event must be reported within 24 hours to the Primary Investigator and Sponsor and within 2 working days hours to Merck Global Safety. (Attn: Worldwide Product Safety; FAX 215 993-1220)

##### 7.2.2 Reporting of Pregnancy and Lactation to the Sponsor and to Merck

Although pregnancy and lactation are not considered adverse events, it is the responsibility of investigators or their designees to report any pregnancy or lactation in a subject (spontaneously reported to them), including the pregnancy of a male subject's female partner that occurs during the trial or within 120 days of completing the trial completing the trial, or 30 days following cessation of treatment if the subject initiates new anticancer therapy, whichever is earlier. All subjects and female partners of male subjects who become pregnant must be followed to the completion/termination of the pregnancy. Pregnancy outcomes of spontaneous abortion, missed abortion, benign hydatidiform mole, blighted ovum, fetal death, intrauterine death, miscarriage and stillbirth must be reported as serious events (Important Medical Events). If the pregnancy continues to term, the outcome (health of infant) must also be reported.

Such events must be reported within 24 hours to the Primary Investigator and Sponsor and within 2 working days to Merck Global Safety. (Attn: Worldwide Product Safety; FAX 215 993-1220)

##### 7.2.3 Immediate Reporting of Adverse Events to the Sponsor and to Merck

###### 7.2.3.1 Serious Adverse Events

A serious adverse event is any adverse event occurring at any dose or during any use of gemcitabine or pembrolizumab that:

- Results in death;
- Is life threatening;
- Results in persistent or significant disability/incapacity;
- Results in or prolongs an existing inpatient hospitalization;
- Is a congenital anomaly/birth defect;

- Is a new cancer (that is not a condition of the study);
- Is associated with an overdose;
- Is another important medical event

Refer to Table 10 for additional details regarding each of the above criteria.

Progression of the cancer under study is not considered an adverse event unless it results in hospitalization or death.

Any serious adverse event, or follow up to a serious adverse event, including death due to any cause other than progression of the cancer under study that occurs to any subject from the time the consent is signed through 90 days following cessation of treatment, or the initiation of new anti-cancer therapy, whichever is earlier, whether or not related to gemcitabine or pembrolizumab, must be reported within 24 hours to the Sponsor and within 2 working days to Merck Global Safety.

Non-serious Events of Clinical Interest will be forwarded to Merck Global Safety and will be handled in the same manner as SAEs.

Additionally, any serious adverse event, considered by an investigator who is a qualified physician to be related to gemcitabine or pembrolizumab that is brought to the attention of the investigator at any time outside of the time period specified in the previous paragraph also must be reported immediately to the Sponsor and to Merck.

An unanticipated event that is serious and definitely or probably caused by the study treatment (drugs or device) will be reported to the IRB in accordance with their guidelines and within their timelines. Collaborating sites must comply with all local IRB requirements and copies of all IRB approvals from these sites must be provided by the site's clinical research staff to the PH&S Regional Research Office.

**SAE reports and any other relevant safety information are to be forwarded to the Merck Global Safety facsimile number: +1-215-993-1220**

A copy of all 15 Day Reports and Annual Progress Reports is submitted as required by FDA, European Union (EU), Pharmaceutical and Medical Devices agency (PMDA) or other local regulators. Investigators will cross reference this submission according to local regulations to the Merck Investigational Compound Number (IND, CSA, etc.) at the time of submission. Additionally investigators will submit a copy of these reports to Merck & Co., Inc. (Attn: Worldwide Product Safety; FAX 215 993-1220) at the time of submission to FDA.

All subjects with serious adverse events must be followed up for outcome.

###### 7.2.3.2 Events of Clinical Interest

Selected non-serious and serious adverse events are also known as Events of Clinical Interest (ECI) and must be recorded as such on the Adverse Event case report forms/worksheets and reported within 24 hours to the Sponsor and within 2 working days to Merck Global Safety. (Attn: Worldwide Product Safety; FAX 215 993-1220)

Events of clinical interest for this trial include:

1. an overdose of Merck product, as defined in Section 7.2.1 - Definition of an Overdose for This Protocol and Reporting of Overdose to the Sponsor, that is not associated with clinical symptoms or abnormal laboratory results.

2. an elevated AST or ALT lab value that is greater than or equal to 3X the upper limit of normal and an elevated total bilirubin lab value that is greater than or equal to 2X the upper limit of normal and, at the same time, an alkaline phosphatase lab value that is less than 2X the upper limit of normal, as determined by way of protocol-specified laboratory testing or unscheduled laboratory testing.*

> <u>*Note:</u> These criteria are based upon available regulatory guidance documents. The purpose of the criteria is to specify a threshold of abnormal hepatic tests that may require an additional evaluation for an underlying etiology. The trial site guidance for assessment and follow up of these criteria can be found in the Investigator Trial File Binder (or equivalent).

3. In the event a subject develops any of the following AEs, a detailed narrative of the event should be reported as an ECI to the Sponsor within 24 hours and to Merck Global Safety within 2 working days of the event:
    a. Grade ≥ 3 diarrhea
    b. Grade ≥ 3 colitis
    c. Grade ≥ 2 pneumonitis
    d. Grade ≥ 3 hypo- or hyperthyroidism

A separate guidance document has been provided entitled "event of Clinical Interest and Immune-Related Adverse Event Guidance Document." This document provides guidance regarding identification, evaluation and management of ECIs and irAEs. Additional ECIs are identified in this guidance document and also need to be reported to the Sponsor within 24 hours and to Merck Global Safety within 2 working days of the event.

Subjects should be assessed for possible ECIs prior to each dose. Lab results should be evaluated and subjects should be asked for signs and symptoms suggestive of an immune-related event. Subjects who develop an ECI thought to be immune-related should have additional testing to rule out other etiologic causes. If lab results or symptoms indicate a possible immune-related ECI, then additional testing should be performed to rule out other etiologic causes. If no other cause is found, then it is assumed to be immune-related.

ECIs that occur in any subject from the date of first dose through 90 days following cessation of treatment, or the initiation of a new anticancer therapy, whichever is earlier, whether or not related to the Merck's product, must be reported within 24 hours to the Sponsor and to Merck Global Safety within 2 working days.

###### 7.2.4 Evaluating Adverse Events

An investigator who is a qualified physician will evaluate all adverse events according to the NCI Common Terminology for Adverse Events (CTCAE), version 4.0 (See Section 12.2). Any adverse event which changes CTCAE grade over the course of a given episode will have each change of grade recorded on the adverse event case report forms/worksheets.

All adverse events regardless of CTCAE grade must also be evaluated for seriousness.

Table 10 Evaluating Adverse Events

An investigator who is a qualified physician, will evaluate all adverse events as to:

| | | |
|---|---|---|
| **V4.0 CTCAE Grading** | **Grade 1** | **Mild; asymptomatic or mid symptoms; clinical or diagnostic observations only; intervention not indicated.** |
| | **Grade 2** | **Moderate; minimal, local or noninvasive intervention indicated; limiting age-appropriate instrumental ADL.** |
| | **Grade 3** | **Severe or medically significant but not immediately life-threatening; hospitalization or prolongation or hospitalization indicated; disabling; limiting self-care ADL.** |
| | **Grade 4** | **Life threatening consequences; urgent intervention indicated.** |
| | **Grade 5** | **Death related to AE** |
| **Seriousness** | A serious adverse event is any adverse event occurring at any dose or during any use of Merck product that: | |
| | †**Results in death**; or | |
| | †**Is life threatening; or** places the subject, in the view of the investigator, at immediate risk of death from the event as it occurred (Note: This does not include an adverse event that, had it occurred in a more severe form, might have caused death.); or | |
| | †**Results in a persistent or significant disability/incapacity** (substantial disruption of one's ability to conduct normal life functions); or | |
| | †**Results in or prolongs an existing inpatient hospitalization** (hospitalization is defined as an inpatient admission, regardless of length of stay, even if the hospitalization is a precautionary measure for continued observation. (Note: Hospitalization [including hospitalization for an elective procedure] for a preexisting condition which has not worsened does not constitute a serious adverse event.); or | |
| | †**Is a congenital anomaly/birth defect** (in offspring of subject taking the product regardless of time to diagnosis);or | |
| | **Is a** new **cancer;** (that is not a condition of the study) **or** | |
| | **Is an overdose** (whether accidental or intentional). Any adverse event associated with an overdose is considered a serious adverse event. An overdose that is not associated with an adverse event is considered a non-serious event of clinical interest and must be reported within 24 hours. | |
| | **Other important medical events** that may not result in death, not be life threatening, or not require hospitalization may be considered a serious adverse event when, based upon appropriate medical judgment, the event may jeopardize the subject and may require medical or surgical intervention to prevent one of the outcomes listed previously (designated above by a †). | |
| **Duration** | Record the start and stop dates of the adverse event. If less than 1 day, indicate the appropriate length of time and units | |
| **Action taken** | Did the adverse event cause the Merck product to be discontinued? | |
| **Relationship to test drug** | Did the Merck product cause the adverse event? The determination of the likelihood that the Merck product caused the adverse event will be provided by an investigator who is a qualified physician. The investigator's signed/dated initials on the source document or worksheet that supports the causality noted on the AE form, ensures that a medically qualified assessment of causality was done. This initialed document must be retained for the required regulatory time frame. The criteria below are intended as reference guidelines to assist the investigator in assessing the likelihood of a relationship between the test drug and the adverse event based upon the available information.<br>**The following components are to be used to assess the relationship between the Merck product and the AE**; the greater the correlation with the components and their respective elements (in number and/or intensity), the more likely the Merck product caused the adverse event (AE): | |
| | **Exposure** | Is there evidence that the subject was actually exposed to the Merck product such as: reliable history, acceptable compliance assessment (pill count, diary, etc.), expected pharmacologic effect, or measurement of drug/metabolite in bodily specimen? |
| | **Time Course** | Did the AE follow in a reasonable temporal sequence from administration of the Merck product?<br>Is the time of onset of the AE compatible with a drug-induced effect (applies to trials with investigational medicinal product)? |
| | **Likely Cause** | Is the AE not reasonably explained by another etiology such as underlying disease, other drug(s)/vaccine(s), or other host or environmental factors |

<table>
<tr><td rowspan="3">Relationship to Merck product (continued)</td><td colspan="2">The following components are to be used to assess the relationship between the test drug and the AE: (continued)</td></tr>
<tr><td>Dechallenge</td><td>Was the Merck product discontinued or dose/exposure/frequency reduced?<br>If yes, did the AE resolve or improve?<br>If yes, this is a positive dechallenge. If no, this is a negative dechallenge.<br>(Note: This criterion is not applicable if: (1) the AE resulted in death or permanent disability; (2) the AE resolved/improved despite continuation of the Merck product; or (3) the trial is a single-dose drug trial); or (4) Merck product(s) is/are only used one time.)</td></tr>
<tr><td>Rechallenge</td><td>Was the subject re-exposed to the Merck product in this study?<br>If yes, did the AE recur or worsen?<br>If yes, this is a positive rechallenge. If no, this is a negative rechallenge.<br>(Note: This criterion is not applicable if: (1) the initial AE resulted in death or permanent disability, or (2) the trial is a single-dose drug trial); or (3) Merck product(s) is/are used only one time).<br>NOTE: IF A RECHALLENGE IS PLANNED FOR AN ADVERSE EVENT WHICH WAS SERIOUS AND WHICH MAY HAVE BEEN CAUSED BY THE MERCK PRODUCT, OR IF REEXPOSURE TO THE MERCK PRODUCT POSES ADDITIONAL POTENTIAL SIGNIFICANT RISK TO THE SUBJECT, THEN THE RECHALLENGE MUST BE APPROVED IN ADVANCE BY THE U.S. CLINICAL MONITOR AS PER DOSE MODIFICATION GUIDELINES IN THE PROTOCOL.</td></tr>
<tr><td></td><td>Consistency with Trial Treatment Profile</td><td>Is the clinical/pathological presentation of the AE consistent with previous knowledge regarding the Merck product or drug class pharmacology or toxicology?</td></tr>
<tr><td colspan="3">The assessment of relationship will be reported on the case report forms /worksheets by an investigator who is a qualified physician according to his/her best clinical judgment, including consideration of the above elements.</td></tr>
<tr><td colspan="2">Record one of the following</td><td>Use the following scale of criteria as guidance (not all criteria must be present to be indicative of a Merck product relationship).</td></tr>
<tr><td colspan="2">Yes, there is a reasonable possibility of Merck product relationship.</td><td>There is evidence of exposure to the Merck product. The temporal sequence of the AE onset relative to the administration of the Merck product is reasonable. The AE is more likely explained by the Merck product than by another cause.</td></tr>
<tr><td colspan="2">No, there is not a reasonable possibility Merck product relationship</td><td>Subject did not receive the Merck product OR temporal sequence of the AE onset relative to administration of the Merck product is not reasonable OR there is another obvious cause of the AE. (Also entered for a subject with overdose without an associated AE.)</td></tr>
</table>

##### 7.2.5 Sponsor Responsibility for Reporting Adverse Events

All Adverse Events will be reported to regulatory authorities, IRB/IECs and investigators in accordance with all applicable global laws and regulations.

#### 8.0 STATISTICAL ANALYSIS PLAN

##### 8.1 Sample Size

The phase I de-escalation portion of the study described in section 5 involves a design with dose cohort size of 6, toxicity evaluated after each 2 enrollments, and only allowing de-escalation. At completion of phase I, data will be collected for at least 6 weeks of follow-up on 6 patients at the MTD or maximum doses studied (200 mg pembrolizumab plus 1250 mg/$m^2$ gemcitabine) if MTD was not reached. In phase II, an additional 10 patients will be enrolled at the final dose from phase I for a total of N=16 subjects. With 24 mo of follow-up, N=16 provides 80% power to detect a difference between median survival times of 3 mo in a historical control compared to 9 mo in the current study, assuming exponential survival distributions and using a 2-sided test at alpha=.05 level of significance. The historical control is based on estimates of median survival times seen in the literature ranging from 2.5-4.25 mo. Total number of patients enrolled will range from 16 to 46 ( (3 gemcitabine doses x 2 pembrolizumab doses x 6/cohort) + 10 in phase II). Sample size was calculated using SAS 9.3 (SAS Institute Inc., Cary, NC).

##### 8.2 Statistical Analysis Plan Summary

**Primary outcome**

- Frequencies of toxicities

**Secondary outcomes**

- Progression-free survival (PFS)
- Overall survival (OS)
- Response rate (CR, PR, SD, PD)

**Exploratory analyses**

- Association between immune response (serial protein array measures from peripheral blood samples) and response rate and survival (progression-free and overall survival) over time, assessed as retrospective analysis.
- Association between immune response time(s) of interest and PFS and OS, assessed as retrospective analysis.

- Association between baseline PD-L1 expression in tumor tissue and PFS, OS, and Best overall response (CR+PR+SD vs PD), assessed as retrospective analysis.

- Association between baseline tumor tissue immune score and PFS, OS, and Best overall response, assessed as retrospective analysis.

#### 8.3 Statistical Analysis Plan

*Safety and feasibility*

If phase I is completed with 0-1 DLTs at the final dose tested, the combination therapy will be considered feasible. Safety will be summarized by listing frequencies of all toxicities at all doses after phase I. Additional safety and other analyses will be performed as needed over follow-up. Analyses will be done for each dose cohort separately.

*Survival analysis*

PFS will be analyzed by modeling time to the first of death or progression. OS will be analyzed by modeling time to death. Patients alive at last date of follow-up will be censored. Both PFS and OS will be described with Kaplan-Meier survival curves. Confidence intervals of median survival times of patients in this study, from both phases I and II, treated with the phase II dose of pembrolizumab and gemcitabine combination therapy will be calculated to see whether they are greater than 3 months, the median survival times from historical controls of patients with previously-treated advanced NSCLC that received gemcitabine single agent therapy.

*Response*

Frequencies of tumor response levels (CR, PR, SD, PD) will be listed for each time point.

*Exploratory analyses*

All exploratory analyses will be performed as retrospective analysis.

Immune response as measured by serial protein array measures from peripheral blood samples at baseline and across follow-up times will be described with means and standard deviations. If within subject changes from baseline to particular follow-up time are observed using paired t-tests, association with outcome Best overall response will be modeled using logistic regression with baseline and change since baseline as independent variables.

Cox proportional hazards regression of PFS and OS will be used to assess univariate effects of baseline tumor PD-L1 expression and tumor immune score measures. Multivariate survival Cox proportional hazards models will be used to test association of PD-L1 and immune score measures while controlling for each other on PFS and OS.

T-tests, or wilcoxon rank-sum tests if nonparametrics are needed, will be used to test the effects of the same baseline PD-L1 expression and immune score measures on best overall response. Log transformations will be used as needed to meet model assumptions.

### 9.0 LABELING, PACKAGING, STORAGE AND RETURN OF CLINICAL SUPPLIES

#### 9.1 Investigational Product

The investigator shall take responsibility for and shall take all steps to maintain appropriate records and ensure appropriate supply, storage, handling, distribution and usage of investigational product in accordance with the protocol and any applicable laws and regulations.

**Gemcitabine** will be obtained as commercially-available product. Refer to the Gemcitabine label for full information regarding product warnings.

**How Supplied**

Gemzar (gemcitabine for injection, USP), is available in sterile single-use vials individually packaged in a carton containing: 200 mg white to off-white, lyophilized powder in a 10-mL size sterile single-use vial – NDC 0002-7501-01 (No. 7501)

1 g white to off-white, lyophilized powder in a 50-mL size sterile single-use vial – NDC 0002-7502-01 (No. 7502)

**Storage and Handling**

Unopened vials of Gemzar are stable until the expiration date indicated on the package when stored at controlled room temperature 20° to 25°C (68° to 77°F) and that allows for excursions between 15° and 30°C (59° and 86°F)

**Pembrolizumab** will be supplied by Merck.

**How Supplied**

Pembrolizumab is provided as a white to off white lyophilized powder (50 mg/vial) or as a liquid solution (100 mg/vial) in Type I glass vials intended for single use only. Pembrolizumab Powder for Solution for Infusion, 50 mg/vial, is reconstituted with sterile water for injection prior to use. Pembrolizumab is formulated with L-histidine as buffering agent, polysorbate 80 as surfactant, sucrose as stabilizer/tonicity modifier, and hydrochloric acid (HCl) and/or sodium hydroxide (NaOH) for pH adjustment (if necessary).

**Storage and Handling**

The drug product is stored as a stable lyophilized powder or liquid solution under refrigerated conditions (2°C - 8°C).

The product after reconstitution with sterile water for injection and the liquid drug product is a clear to opalescent solution which may contain extraneous and proteinaceous particulates. The reconstituted product and liquid product is intended for IV administration. The reconstituted drug product solution and liquid drug product can be further diluted with normal saline in IV containers made of polyvinyl chloride (PVC) or non-PVC material. Reconstituted vials should be immediately used to prepare the infusion solution in the IV bag and the infusion solution should be immediately administered. If not used immediately, vials and/or IV bags may be stored at 2-8 °C for up to a cumulative time of 20 hours. If refrigerated, the vials and/or IV bags should be allowed to equilibrate to room temperature prior to subsequent use. Pembrolizumab solutions may be stored at room temperature for a cumulative time of up to 4 hours. This includes room temperature storage of reconstituted drug product solution and liquid drug product in vials, room temperature storage of infusion solution in the IV bag and the duration of infusion.

Please refer to the pembrolizumab Investigator's Brochure for additional product information.

#### 9.2 Packaging and Labeling Information

Clinical supplies will be affixed with a clinical label in accordance with regulatory requirements.

#### 9.3 Clinical Supplies Disclosure

This trial is open-label; therefore, the subject, the trial site personnel, the Sponsor and/or designee are not blinded to treatment. Drug identity (name, strength) is included in the label text; random code/disclosure envelopes or lists are not provided.

#### 9.4 Storage and Handling Requirements

Clinical supplies must be stored in a secure, limited-access location under the storage conditions specified on the label.

Receipt and dispensing of trial medication must be recorded by an authorized person at the trial site.

Clinical supplies may not be used for any purpose other than that stated in the protocol.

#### 9.5 Returns and Reconciliation

The investigator is responsible for keeping accurate records of the clinical supplies received from Merck or designee, the amount dispensed to and returned by the subjects and the amount remaining at the conclusion of the trial.

Upon completion or termination of the study, all unused and/or partially used investigational product will be destroyed at the site per institutional policy. It is the Investigator's responsibility to arrange for disposal of all empty containers, provided that procedures for proper disposal have been established according to applicable federal, state, local and institutional guidelines and procedures, and provided that appropriate records of disposal are kept.

### 10.0 ADMINISTRATIVE AND REGULATORY DETAILS

#### 10.1 Protocol Modifications and Amendments

All modifications or amendments to the protocol or informed consent document must be approved by the Principal Investigator and submitted to the PH&S IRB for review and approval. All modifications and amendments will be documented with a new version number and date. All changes to the informed consent document will include the date of the revision on the form.

No changes will be implemented until IRB approval is obtained except when a potential threat to patient safety exists.

The IRB will be notified of any significant deviations from the approved protocol. Documentation of all IRB correspondence will be maintained in the central regulatory file.

#### 10.2 Continuing Review and Final Reports

An annual progress report (continuing review) will be submitted to the PH&S IRB for the duration of the study. A final report to the PH&S IRB will be submitted at the summation of the study.

#### 10.3 Record Retention

According to 21 CFR 312.62(c), the investigator shall retain required records for a period of 2 years following the date a marketing application is approved for the drug for the indication for which it is being investigated. If no application is to be filed or if the application is not approved for such indication, the investigator shall retain these records until 2 years after the investigation is discontinued or the IND is withdrawn and the FDA is notified.

The investigator must retain protocols, amendments, IRB/IBC approvals, completed, signed, dated consent forms, patient source documents, case report forms, quality monitoring reports, drug accountability records and all documents of any nature regarding the study or patients enrolled. All records will be maintained under restricted access by the Clinical Trials Department at Providence Portland Medical Center while the study remains active. Records may be placed in long-term storage after the study is completed. The location of long-term storage will be secure and easily accessed for regulatory purposes.

#### 10.4 Confidentiality

Study patients will be identified by their assigned study number on all forms and biospecimen sample labels. Patient study data will be collected on eCRFs that are maintained in the Velos eResearch CTMS. eResearch is a web-based application that is accessed using a unique username and password (one login per user). Patient study charts are maintained in a keycode access file room within the Clinical Trials Department.

#### 10.5 Compliance with Financial Disclosure Requirements

Any investigator conducting Research at Providence must comply with the Providence COIR Policy and complete a Providence COIR disclosure form. The COIR disclosure form must be completed prior to submitting Research studies through a Providence IRB or non-Providence IRB of record, prior to submitting a grant; regardless of the source of funding.

#### 10.6 Compliance with Trial Registration and Results Posting Requirements

Under the terms of the Food and Drug Administration Modernization Act (FDAMA) and the Food and Drug Administration Amendments Act (FDAAA), the Sponsor of the trial is solely responsible for determining whether the trial and its results are subject to the requirements for submission to the Clinical Trials Data Bank, http://www.clinicaltrials.gov. Information posted will allow subjects to identify potentially appropriate trials for their disease conditions and pursue participation by calling a central contact number for further information on appropriate trial locations and trial site contact information.

#### 10.7 Quality Management System

Quality Assurance (QA) personnel review study monitoring reports and if necessary, determine follow-up actions to resolve significant findings. QA has the authority to request immediate corrective action if significant patient safety issues are identified.

QA will track and trend results from study monitoring reports as well as associated corrective and preventive actions. A QA summary report will be provided to the IRB at the time of continuing review.

QA personnel do not have a direct reporting relationship to the Principal Investigator and are not responsible for enrollment or coordination of care for study participants.

#### 10.8 Data Management

**Study Monitoring**

Study monitoring activities (Quality Control Reviews) are performed by a Contract Research Organization (CRO) or clinical research staff members who have completed specialized training in study monitoring procedures and human subjects' protections. Individuals who perform study monitoring activities do not report to Principal Investigators or research scientists and may not monitor studies for which they have direct responsibility.

Study monitoring activities are conducted regularly and include (but are not limited to) review and verification of the following:

- Eligibility
- Informed Consent process
- Adherence to protocol treatment plan
- Case Report Forms (CRFs)

- Source Documentation
- Adverse Events
- Regulatory Reporting

Results of study monitoring activities will be reported to applicable study personnel, the Clinical Trials Manager and Quality Assurance.

**Data Reporting**

eCRFs will be completed in Velos eResearch Clinical Trials Management System, a part 11 compliant application.

#### 11.0 LIST OF REFERENCES

Confidential ✪ Limited Access

32. Brahmer JR, Horn L, Antonia SJ, et al. Nivolumab in patients with non-small cell lung cancer: Overall survival and long-term safety in a phase 1 trial. J Thoracic Oncol 2013; 8: S365 (abstr).
33. Garon EB, Leighl NB, Rizvi NA, et al. Safety and clinical activity of MK-3475 in previously treated patients with non-small cell lung cancer (NSCLC). Proc Am Soc Clin Oncol 2014; 32: 8020 (abstr).
34. Nowak AK, Lake RA, Marzo AL, et al. Induction of tumor cell apoptosis in vivo increases tumor antigen cross-presentation, cross-priming rather than cross-tolerizing host tumor-specific CD8 T cells. J Immunol 2003; 170: 4905-4913.
35. Nowak AK, Robinson BWS, Lake RA. Synergy between chemotherapy and immunotherapy in the treatment of established murine solid tumors. Cancer Research 2003; 63: 4490-4496.
36. Pages F, Kirilovsky A, Mlecnik B, et al. In situ cytotoxic and memory T cells predict outcome in patients with early-stage colorectal cancer. J Clin Oncol 2009; 27: 5944-5951.
37. Mlecnik B, Tosolini M, Kirilovsky A, et al. Histopathologic-based prognostic factors in colorectal cancers are associated with the state of the local immune reaction. J Clin Oncol 2011; 29: 610-618.
38. Ascierto, PA, Capone M, Urba WJ, Bifilco CB, Botti G, Lugli A, Marincola FM, Ciliberto G, Galon J, Fox BA, The additional facet of immunoscore: immunoprofiling as a possible predictive tool for cancer treatment. J Transl Med, 2013 Mar 11(1):54. PMCID: *PMID:23452415*
39. Galon J, Mlecnik B, Bindea G, Angell HK, Berger A, Lagorce C, Galon J, Mlecnik B, Bindea G, Angell HK, Berger A, Lagorce C, Lugli A, Zlobec I, Hartmann A, Bifulco C, Nagtegaal ID, Palmqvist R, Masucci GV, Botti G, Tatangelo F, Delrio P, Maio M, Laghi L, Grizzi F, Asslaber M, D'Arrigo C, Vidal-Vanaclocha F, Zavadova E, Chouchane L, Ohashi PS, Hafezi-Bakhtiari S, Wouters BG, Roehrl M, Nguyen L, Kawakami Y, Hazama S, Okuno K, Ogino S, Gibbs P, Waring P, Sato N, Torigoe T, Itoh K, Patel PS, Shukla SN, Wang Y, Kopetz S, Sinicrope FA, Scripcariu V, Ascierto PA, Marincola FM, **Fox BA**, Pagès F: Towards the introduction of the Immunoscore in the classification of malignant tumors. J Pathol. 2014 Jan;232(2):199-209. *PMCID: PMC3681569*
40. Suzuki K, Kadota K, Sima CS, et al. Clinical impact of immune microenvironment in stage I lung adenocarcinoma: Tumor interleukin-12 receptor β2, IL-7R, and stromal FoxP3/CD3 ratio are independent predictors of recurrence. J Clin Oncol 2013; 31: 409-498.
41. Graff JN, Puri S, Bifulco C, Fox BA, Beer TM. Sustained Complete Response to CTLA-4 Blockade in a Patient with Metastatic, Castration-Resistant Prostate Cancer. Cancer Immunology Research *Published OnlineFirst February 3, 2014; doi:10.1158/2326-6066.CIR-13-0193*

### 12.0 APPENDICES

#### 12.1 ECOG Performance Status

| Grade | Description |
|---|---|
| 0 | Normal activity. Fully active, able to carry on all pre-disease performance without restriction. |
| 1 | Symptoms, but ambulatory. Restricted in physically strenuous activity, but ambulatory and able to carry out work of a light or sedentary nature (e.g., light housework, office work). |
| 2 | In bed <50% of the time. Ambulatory and capable of all self-care, but unable to carry out any work activities. Up and about more than 50% of waking hours. |
| 3 | In bed >50% of the time. Capable of only limited self-care, confined to bed or chair more than 50% of waking hours. |
| 4 | 100% bedridden. Completely disabled. Cannot carry on any self-care. Totally confined to bed or chair. |
| 5 | Dead. |
| * As published in Am. J. Clin. Oncol.: *Oken, M.M., Creech, R.H., Tormey, D.C., Horton, J., Davis, T.E., McFadden, E.T., Carbone, P.P.: Toxicity And Response Criteria Of The Eastern Cooperative Oncology Group. Am J Clin Oncol 5:649-655, 1982.* The Eastern Cooperative Oncology Group, Robert Comis M.D., Group Chair. | |

#### 12.2 Common Terminology Criteria for Adverse Events V4.0 (CTCAE)

The descriptions and grading scales found in the revised NCI Common Terminology Criteria for Adverse Events (CTCAE) version 4.0 will be utilized for adverse event reporting. (http://ctep.cancer.gov/reporting/ctc.html)

#### 12.3 Response Evaluation Criteria in Solid Tumors (RECIST) 1.1 Criteria for Evaluating Response in Solid Tumors

RECIST version 1.1* will be used in this study for assessment of tumor response. While either CT or MRI may be utilized, as per RECIST 1.1, CT is the preferred imaging technique in this study.

* As published in the European Journal of Cancer:

E.A. Eisenhauer, P. Therasse, J. Bogaerts, L.H. Schwartz, D. Sargent, R. Ford, J. Dancey, S. Arbuck, S. Gwyther, M. Mooney, L. Rubinstein, L. Shankar, L. Dodd, R. Kaplan, D. Lacombe, J. Verweij. New response evaluation criteria in solid tumors: Revised RECIST guideline (version 1.1). Eur J Cancer. 2009 Jan;45(2):228-47.
