## Supplemental Figure 8 for "Provides Apparent Predictive Biomarker in a Phase I/II Study of Pembrolizumab With Gemcitabine in Patients with Previously-Treated Advanced Non-Small Cell Lung Cancer (NSCLC)"

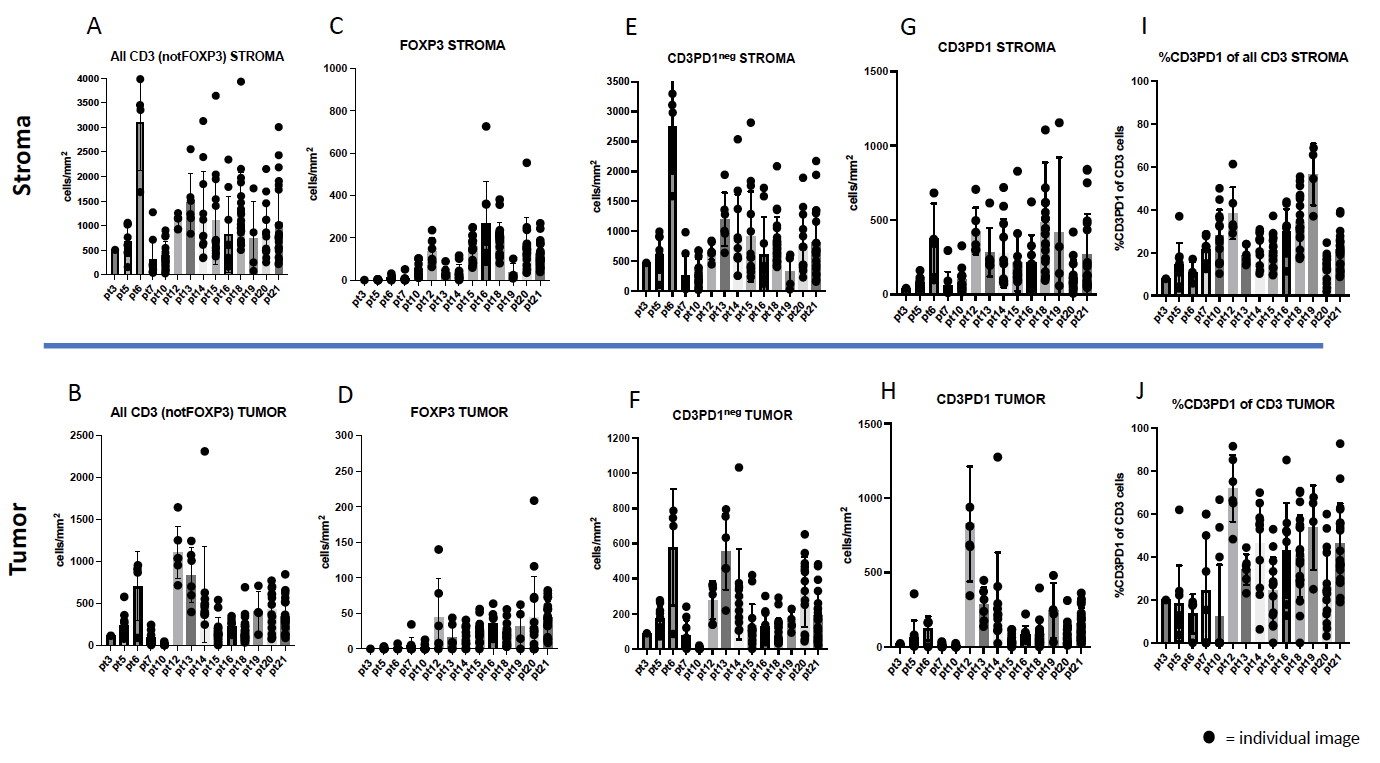

Supplemental Figure 8. *The cell density (cells/mm^2^) of CD3^+^FoxP3^neg^ in the stroma region (A) or tumor region (B); CD3^+^FoxP3^+^ in the stroma region (C) or tumor region (D); CD3^+^PD-1^neg^ in the stroma region (E) or tumor region (F); CD3^+^PD-1^neg^ in the stroma region (G) or tumor region (H) are shown. The percent T cells (CD3^+^) expressing PD-1 in the stroma region (I) and tumor region (J).*
